# From sample to insight: Onsite training and implementing mobile environmental surveillance in sub-Saharan Africa

**DOI:** 10.64898/2026.07.21.26358224

**Authors:** Tam Tran, Jeremy Cook, Dylan Shea, Andrea Bagi, Gro Fonnes, Alan Le Tressoler, Peter Mkama, Eric Lyimo, Hillary Sebukoto, Jackson Claver, Melissa Kabena, Evodie Ngelesi, Placide Mbala, Palpouguini Lompo, Bérenger Kaboré, Sallamaari Siponen, Elisa Salmivirta, Vito Baraka, Marc Christian Tahita, Vivi Maketa Tevuzula, Tarja Pitkänen, Rolf Lood, Adriana Krolicka

## Abstract

Environmental surveillance for human pathogen detection is crucial for safeguarding public health; however, traditional approaches are often time-consuming, equipment-intensive and workflow-complex, delaying actionable public-health decision-making. The integration of a mobile laboratory (ML) with rapid bioinformatic analysis and interactive data visualization offers a novel approach to this surveillance. The ML workflows, with a focus on metagenomic sequencing and qPCR technologies, have been specifically designed to provide real-time information in detecting epidemiological threats in environmental samples. We have adopted a minimalist strategy, avoiding complex, expensive, and energy-intensive equipment that is challenging to maintain and transport, thereby reflecting a more sustainable approach. This study details the activities and outcomes of implementing such a ML in Tanzania. During one-month ML deployment, a total of 83 samples were sequenced using long-read metagenomic sequencing, and 70 samples were analyzed by Biomeme qPCR. The deployment enabled the detection of multiple bacterial, viral, and antimicrobial resistance targets in wastewater, drinking water, surface water, and soil samples providing valuable insights into the population epidemiological information relevant to the prevention of waterborne and zoonotic infections. In addition, the ML deployment provided a rare opportunity for knowledge transfer, actively engaged local researchers, and strengthened local epidemiological capacity. After the deployment, certificates of completion were issued to twenty-five scientists from four African countries (Tanzania, Burkina Faso, Democratic Republic of Congo, and Ethiopia) and post-deployment surveys were sent out to gather their feedback. Our findings highlight the importance of combining targeted qPCR assays with metagenomic sequencing for the proper interpretation, while acknowledging the risk of false-positive detections arising from low-abundance sequencing reads, particularly when using fast taxonomic classification tools such as kraken2. Given the hardware limitations inherent to ML deployments, metagenomic pathogen detection in this context should therefore be regarded primarily as a valuable screening tool rather than a definitive diagnostic method. These observations further underscore the need for more stringent and computationally demanding bioinformatic approaches when assessing high-priority pathogens. Complementary qPCR-based assays conducted within the ML can ensure equivalent robustness to those conducted in conventional laboratory settings. This innovative approach has the potential to significantly enhance detection capabilities and support timely public health responses in resource-limited settings.

## 1 Introduction

Increases in urbanization, global travel, human-wildlife interaction, and climate change are altering the disease emergence landscape ^1–3^, necessitating innovation in disease surveillance programs. Traditional surveillance programs focus on passive, facility-based reporting systems where those experiencing symptoms seek healthcare and wait for a test result ^4,5^. Meanwhile, environmental surveillance offers a proactive approach that tracks community-level pathogens before they reach healthcare settings ^6^. In the context of resource-limited settings, this has been proven as a cost-effective alternative to clinical surveillance due to a high-burden cost of disease diagnosis and treatment for these countries ^7^.

Mobile laboratories (MLs) are portable units that can be deployed quickly to various locations ^8–11^. They are equipped with essential laboratory equipment and supplies necessary for conducting molecular biology analyses such as qPCR and metagenomic sequencing. The advantages of MLs include: (1) Rapid Deployment: MLs can be transported to outbreak sites or areas with suspected contamination. (2) On-Site Testing: Immediate testing reduces the time between sample collection and results, facilitating timely public health responses. In addition to making timely decisions, it also enables analysis where samples cannot be safely stored or transported. (3) Capacity Building: Training local personnel in MLs enhances local expertise in pathogen detection. MLs have been in use for decades as essential tools for infectious disease monitoring and detection, whether as part of research initiatives or outbreak response efforts, with their composition varying based on the specific work they are designed to support and the geographical terrain in which they are deployed ^11^.

However, most MLs are primarily deployed for clinical diagnostic purposes to detect known pathogens in patient samples during acute outbreaks. For example, MLs were established across Eastern Africa (Kenya, Burundi, Rwanda, Tanzania, Uganda, and South Sudan) for the detection and management of epidemic-prone diseases such as Dengue fever, Ebola, and COVID-19 ^12,13^. Similarly, MLs were rapidly deployed in responding to the Ebola virus outbreak in West Africa ^9,10^ and detection of viral hemorrhagic fevers in Johannesburg, South Africa ^8^. MLs were also deployed in other geographical regions beyond sub-Saharan Africa including China, India, Vietnam, Republic of Uzbekistan ^14–18^.

Our previous study has developed flexible and field-deployable modular lab workflows that are tailored to resource-limited settings ^19^. However, for full operational evaluation, onsite deployment of a whole end-to-end process from sampling to analysis is required. To achieve that, the ML in this study was also coupled with a simplified, user-friendly bioinformatic analysis and interactive data visualization through a purpose-built, internet-independent dashboard, allowing actionable interpretation of results in the field. The implementation of the ML lasted the entire month of September 2025 in Tanzania and was combined with training sessions for scientists from Ethiopia, Tanzania, Burkina Faso and the Democratic Republic of the Congo (DRC).

Tanzania was selected as the primary deployment site based on its experience and established operational role being beneficiary of the East African community (EAC) ML network initiative. The National Public Health Laboratory (NPHL) has demonstrated a rapid-response readiness in deploying MLs in several outbreaks’ preparedness. Currently, they still maintain a fully operational ML. In addition, Tanzania hosts the EAC Secretariat and the National Health Laboratory Quality Assurance and Training Centre. This field experience, technical expertise and autonomy, institutional infrastructure capacity made NPHL in Tanzania the strong pilot site for mobile surveillance. As part of the ODIN and ODIN-Mpox consortium, researchers from DRC and Burkina Faso participated in the deployment by contributing previously prepared country-specific sample materials, enabling the ML to be tested across a range of sample provenances ^20^.

The study described here was guided by three primary objectives: (1) deployment and autonomous operation of a modular mobile environmental surveillance laboratory; (2) field generation, analysis, and visualisation of data on priority pathogens from diverse environmental samples using both targeted (qPCR) and untargeted (metagenomic sequencing) approaches; and (3) training and competence building with researchers from partner countries to enable future independent operation.

## 2 Methods

### 2.1 Deployment preparation and mobile lab setup

#### Reagent sourcing

Wherever possible, reagents and consumables were sourced locally in Tanzania and through regional African suppliers. This strategy was adopted not only to support local procurement and strengthen sustainable supply chains for future deployments, but also to establish and maintain relationships with reliable regional distributors of critical laboratory reagents. When local or regional sourcing was not feasible, non-temperature-sensitive reagents and consumables were procured in Norway and transported or shipped to Tanzania ahead of deployment. Temperature-sensitive reagents such as molecular enzymes and master mixes were preferentially sourced through distributors in Tanzania. Initial comparisons were performed between African-sourced reagents and those used in Norwegian validations to verify comparable performance ^19^. The greatest procurement challenge was the acquisition of qPCR assays and oligonucleotides (primers and probes), which were not readily available through regional suppliers and therefore had to be imported.

#### Physical deployment

Required components: large inflatable tent, small collapsible tent, gas generator, mobile fridge and freezer, collapsible tables, equipment and consumables packed into ∼4 aluminium Zargas boxes. - Full assembly/disassembly time: ∼3 hours. - All components fit within two SUVs/vans — enabling rapid deployment to remote settings. - Power: for the majority of the one-month deployment, the ML operated using a gas-powered generator.

The layout of the ML is illustrated in Figure 1 and Figure 2. The ML consisted of semi-protected spaces dedicated to the various stages of sample processing and data acquisition.

**Figure 1:** Mobile environmental surveillance laboratory deployment in Tanzania (A) Overview of the three-tent deployment configuration, including laboratory workspaces and power generation infrastructure. (B) Workstation used for environmental sample processing and nucleic acid extraction. This was set up in Tent 1 to avoid contamination in the downstream process which took place in Tent 2. Tent 3 was used for operational support, including extra storage space and drinking water supply for the team. *Photographs removed from the preprint version in accordance with the preprint server policy on identifiable individuals. Images are available from the corresponding author upon reasonable request*.

**Figure 2:** Downstream process in Tent 2: A) PCR and DNA library preparation workstation (B) Nanopore sequencing and bioinformatics station (C) PCR and qPCR thermocyclers (D) Cold storage (refrigerator and freezer) (E) Qubit fluorometer *Photographs removed from the preprint version in accordance with the preprint policy on identifiable individuals. Images are available from the corresponding author upon reasonable request*.

### 2.2 Sampling campaign, sample pre-processing and nucleic acid extraction

#### 2.2.1 Sampling campaign and metadata collection

Samples were collected at various locations in three countries (Table S1). The sampling work was a joint effort among participating partners in the ODIN and ODIN-Mpox projects. A comprehensive list of samples with their associated information was documented in the metadata file. This metadata file was also subsequently used to run the bioinformatic pipeline.

Samples entered the mobile laboratory workflow at different stages depending on their provenance (Figure 3): - Pathway A — ML-collected samples (full pipeline): Collected on-site by the mobile laboratory team during the Tanzania deployment, these samples passed through every module: collection, filtration, nucleic acid extraction, library preparation (or qPCR), sequencing, and bioinformatic analysis. - Pathway B — Partner-collected samples (extracted on-site): Previously collected by Tanzanian partner institutions (NIMR) and stored frozen, these samples were extracted and analysed in the mobile laboratory, bypassing the collection/filtration step. - Pathway C — Partner nucleic acid extracts (analysis only): Purified DNA/RNA provided by partner institutions in the DRC and Burkina Faso, these entered the workflow at the library preparation or qPCR stage, allowing the mobile laboratory to be tested on material from all three countries even where on-site sample collection was not possible.

**Figure 3:**
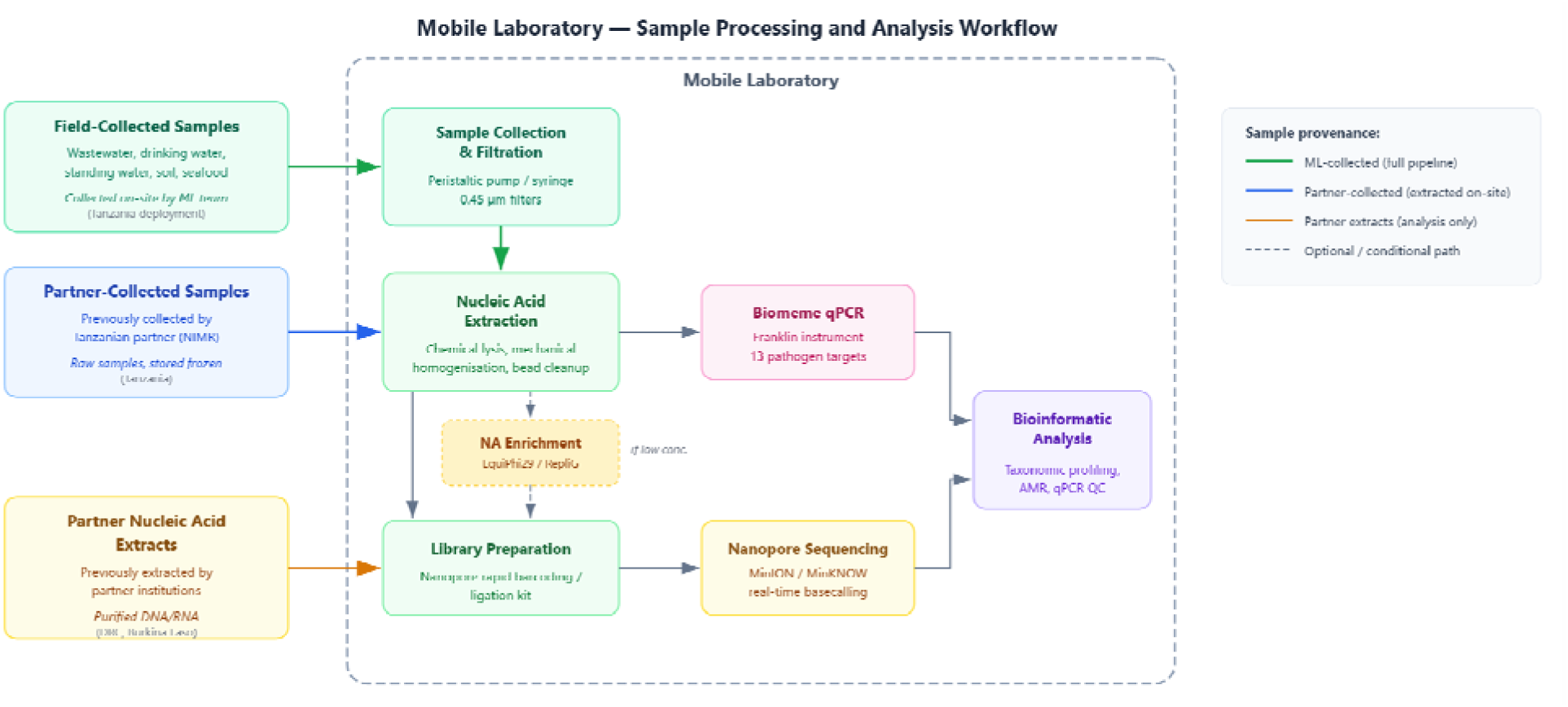
Workflow diagram showing the three sample processing pathways into the mobile laboratory.

This tiered approach demonstrated that the mobile laboratory could operate across the full processing chain while also functioning as an analysis endpoint for material processed elsewhere. The complete sample processing and analysis workflow is shown in Figure 3.

##### Onsite sample Collection

During the ML deployment, sampling was conducted across 12 sites in Dar es Salaam. Multiple environmental matrices were collected at some sites, yielding a total of 30 sampling events: wastewater (n=7), surface water (n=9), drinking water (n=8), and soil (n=6; Table S1) (Figure 4).

**Figure 4.**
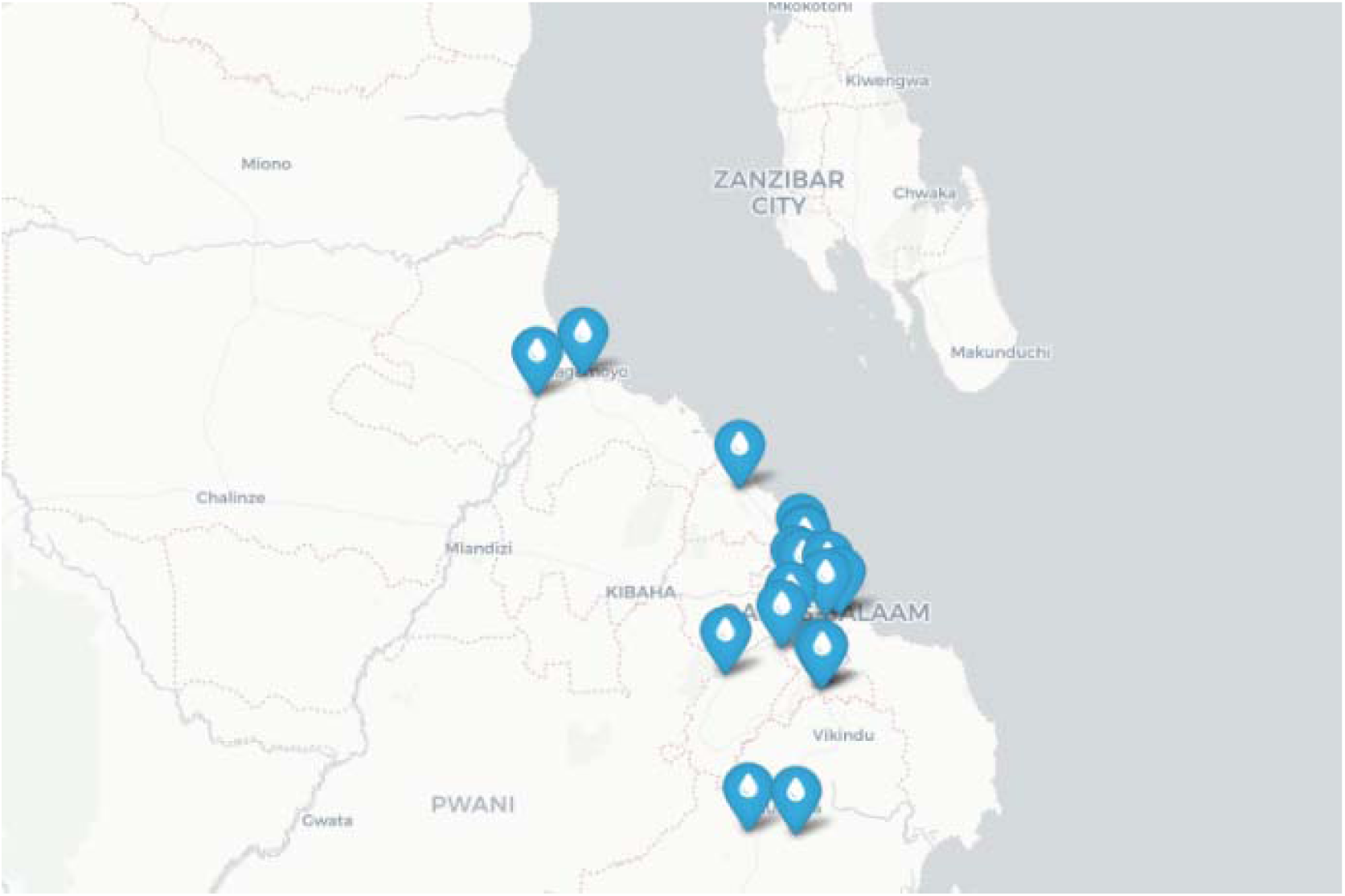
Sampling sites in Dar es Salaam, Tanzania

###### Wastewater

Wastewater samples were collected from seven sampling sites during the ML deployment. Wastewater samples were collected by dipping a beaker affixed to a sampling pole into the wastewater effluent reservoir and transferring water to a sterilized 500 mL collection vessel for transportation to the ML for filtration. Collection vessels containing wastewater were individually bagged and stored with ice packs for transportation to the mobile lab. All collection equipment and collection vessels were sterilized with 10% bleach between collections.

###### Surface water

Surface water sites encompassed natural water sources such as rivers and reservoirs that were used by local populations for washing and in some cases drinking water. Unlike drinking water sites, these locations did not represent dedicated drinking water sources and as such were not subject to treatment or chlorination. Water samples were collected from nine sampling sites during the deployment. At each site, 5 L of water was collected and transferred to a sterilized collection vessel.

###### Drinking water

Drinking water samples were collected from spigots or taps used by local populations. Water samples were collected from eight drinking water sites during the ML deployment. At each site, 5 L of water was collected and transferred to a sterilized collection vessel and kept cool during transportation to the ML for filtration.

###### Soil

Sites where soil could be contaminated by wastewater or washing water runoff from local populations (e.g. markets, dried canals) were selected for soil sample collection. Soil samples were collected from six sampling sites during the deployment. At each sampling site, ∼ 5 g of surface (0 - 1 cm) soil was collected using a disposable sampling spoon from 3 sub-sampling locations within a 1 m radius and combined into a labeled 15 mL collection tube. Tubes containing soil were kept cool during transportation to the mobile lab and stored at -20°C prior to nucleic acid extraction. Soil sampling was conducted in Tanzania only; no soil samples were collected in Burkina Faso or DRC.

Metadata capture was integrated into sample collection and processing workflows; the full metadata model, validation approach, and downstream integration are described in Section 2.3.

Automated pre-run validation was applied before each pipeline execution to verify consistency between metadata and run outputs (Section 2.3).

#### 2.2.2 Filtration

Water samples were filtered upon returning to the ML from collection sites.

Mean filtration volumes varied substantially among drinking water (1500 mL ± 0 mL), surface water (804 mL ± 544 mL), and wastewater (178 mL ± 181 mL) samples due to differences in sample turbidity.

Appropriate personal protective equipment including safety glasses, masks, gloves, and single-use lab coats was used during wastewater filtration. Filter holders, tubing, and forceps were cleaned with 10% bleach between samples. Sample-specific filtration methods are briefly described below but see our previous study for a detailed description of sample-specific filtration methodologies ^19^.

##### Surface Water and Drinking Water

Surface and drinking water samples were filtered through 0.22 mm CN filters atop a polycarbonate filter holder (Sartorius, Germany), with a target volume of up to 1500 mL per filtration replicate. A peristaltic pump (Watson Marlow, UK) was transported from Norway because no suitable local supplier could be identified despite extensive effort putting into this. This pump was used to filter water samples at a rate of 0.2 L/minute, and filtration was terminated for each filtration replicate once 1500 mL had been filtered or once the flow rate was observed to decrease due to filter clogging. When the target filtration volume was not achieved, the final filtration volume for each replicate was recorded. After terminating filtration, each filter was collected and transferred to a labeled 2 mL tube and stored at -20°C prior to nucleic acid extraction.

##### Wastewater

Wastewater samples were filtered immediately upon returning to the ML. Briefly, wastewater samples were loaded into a 50 mL syringe and filtered through a 0.22 mm cellulose nitrate filter (Sartorius, Germany) atop a 25 mm syringe filter holder (Sartorius, Germany). When filtration slowed due to clogging, the used filter was removed and placed into a labeled 2 mL tube, and a new filter was placed in the filter holder to continue filtration. To account for sample turbidity and maximize filtered volume, a total of 3 filters were used for each wastewater sample. All filters corresponding to a given sample were combined in the same 2 mL tube and stored at -20°C prior to nucleic acid extraction.

#### 2.2.3 Nucleic acid extraction

During the ML deployment, nucleic acids (NAs) were extracted from 50 samples. Of the 50 samples extracted in the ML, 30 were collected during the deployment period and 20 were previously collected by Tanzanian collaborators during routine environmental surveys as part of the ODIN project (Table S1).

Nucleic acid extractions were performed in the mobile laboratory using a custom-designed protocol that minimized the need for large, power-intensive lab equipment. Our previous study described the mobile NA extraction protocol in detail ^19^. Briefly, samples were lysed using a combination of chemical lysis buffer (RLT Buffer, Qiagen with dithiothreitol, DTT) and physical disruption (vortex). Following lysis, samples were mechanically homogenized by pipetting up and down through a 20G needle attached to a syringe. Cellular debris was removed by filtering samples through a Millex-GV filter then subjected to a HighPrep PCR-DX magnetic bead-based purification with a two-step binding and wash protocol. Purified nucleic acids were stored at −20°C prior to downstream molecular analysis. NA extracts were aliquoted to facilitate analysis via qPCR and sequencing molecular workflows.

DNA concentrations were measured using the Qubit dsDNA High Sensitivity Assay Kit (Thermo Fisher, USA) on a Qubit fluorometer. For samples subjected to sequencing workflows, NA concentrations were also measured after library preparation to ensure equimolar concentrations of each sample were included on a single sequencing run.

### 2.3 Metadata management and validation

A data dictionary was maintained for the workbook to define field names, allowed values, code mappings, and required versus optional entries. This dictionary reduced ambiguity across teams and countries, and was used during training to standardize data entry and interpretation before pipeline execution and dashboard review.

The workbook (Figure 5) was organized into linked components (sites, samples, nanopore, biomeme) with explicit relational keys. The sites tab stored location-level descriptors and coordinates; samples linked each collected specimen to a site and sampling context; nanopore linked sequencing outputs to sample records; and biomeme linked qPCR runs to the same sample records. This relational structure enabled integration of metagenomics, AMR, and qPCR outputs without manual relabeling.

**Figure 5:**
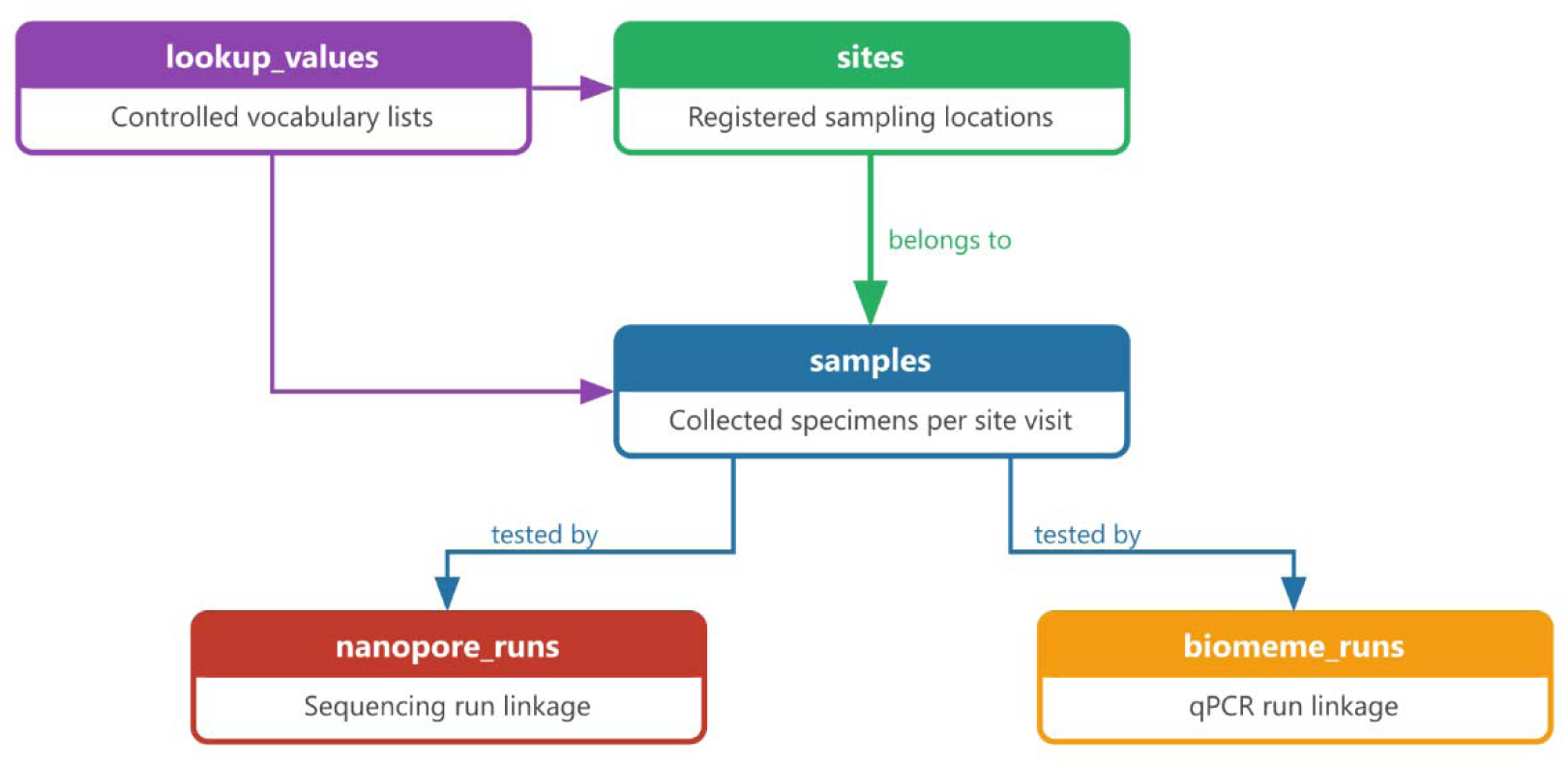
ODIN Metadata Workbook — Component Relationships. Site, sample, sequencing, and qPCR records were linked by relational keys in the form of unique sample identifiers. The sites tab records registered sampling locations; samples linked each collected specimen to a site; nanopore_runs and biomeme_runs linked sequencing and qPCR run outputs to the same sample records. Controlled vocabulary lists (lookup_values) constrained field entries across all components to ensure consistent naming across partners and countries.

### 2.4 Biomeme quantitative PCR (qPCR)

NA aliquots were used for qPCR directly after purification without pre-enrichment as previously described ^19^. Briefly, a master mix containing (per reaction) 10 mL Luna Universal Probe qPCR Master Mix, along with forward and reverse primers (400 nM) and probe (200 nM) for each target assay was prepared on a dedicated nucleic-acid free bench. The prepared master mix was vortexed to mix thoroughly and 18 µL was added to each of nine GoStrip wells. Two GoStrip wells were reserved for no-template control (NTC) reactions each receiving 2 mL of water in place of DNA template. GoStrips^TM^ containing master mix were then transferred to a separate bench for the addition of DNA template. qPCR assays were configured for priority pathogens: Generic Orthopox, Generic Mpox, MPXV clade IB, MPXV clade I, V. cholerae (ctxA), S. typhi (STadh), K. pneumoniae (Khe), and E. coli (uidA). Samples were tested in duplicate, with one GoStrip well reserved for gBlocks positive controls for each target assay. Sealed GoStrips were placed in the Biomeme thermocycler and run on the thermocycling program: 95 °C for 6 minutes, 45 cycles of denaturation at 95 °C for 5 seconds, annealing/extension at 60 °C for 20 seconds. Run progress was monitored using a Bluetooth connected smartphone and, upon completion, data were exported to one of the dedicated laptops for processing and visualization.

### 2.5 Metagenomic Sequencing

The metagenomic sequencing workflow was employed to assess samples for the presence of priority pathogen species and AMR genes in environmental samples. Prior to sequencing, samples were subjected to enrichment via multiple displacement amplification (MDA) using EquiPhi29 DNA polymerase and 5’-phosphorylated random hexamers (Thermofisher, USA). MDA products were purified with HighPrep PCR-DX MagBeads (MagBio, USA) and treated with T7 endonuclease. Sequencing libraries were prepared using the ONT Native Barcoding Kit (ONT, UK) where sample-specific barcodes were ligated prior to equimolar pooling. Pooled libraries were loaded onto ONT flow cells and sequenced on a MinION device using MinKNOW v23.07.15. Reads were basecalled and demultiplexed following ONT’s standard pipeline and reads were filtered using recommended quality thresholds (Q-score ³ 7; minimum length 200 bp). Sequence data were transferred to one of the dedicated laptops for downstream bioinformatic analyses. All raw sequencing reads generated during this study have been deposited in the European Nucleotide Archive (ENA) under BioProject accession PRJEB118616 and are publicly available.

### 2.6 MPXV tiling amplicon sequencing

Twenty-four extracted environmental DNA samples across three countries and five MPXV DNA samples from confirmed clinical cases in DRC, all of which were anonymized prior to molecular analysis, were amplified using a tiling amplicon PCR approach as described in our previous study. ^19^. Briefly, two primer pools, each containing 24 primers, were used to generate overlapping 24 amplicons (12 amplicons per pool) across the MPXV genome. PCR reactions (25 mL) were set up separately for each of the two primer pools using LongAmpTM Hot Start Taq 2X Master Mix (New England Biolabs, USA). PCR reactions were performed under thermocycling conditions: 94 °C for 30 s; 25 cycles of 94 °C for 30 s, 55 °C for 40 s, 65 °C for 9 mins; followed by a final extension at 65 °C for 10 mins. Negative control was included in each run using water as a template. PCR products from both primer pools were combined for each sample and purified using HighPrep PCR-DX MagBeads (MagBio, USA) and eluted in 15 mL EB buffer (Qiagen, Germany). Sequencing libraries were prepared using the Rapid Barcoding Kit (ONT, UK) and sequenced on a R10.4.1 flow cell with a MinION device. Reads were base-called and filtered in high-accuracy mode (Q-score 7; minimum length 200 bp) using MinKNOW v23.07.15. Sequence data were analyzed as described in section 2.9. In some cases, a second-round PCR was performed using first-round PCR product as a template.

### 2.7 Laptop configuration, bioinformatic pipelines and databases for onsite data analysis and visualization

Two ODIN laptops were purchased for use during the deployment. These laptops were used for performing sequencing runs, executing bioinformatic pipelines, and visualizing data. The ODIN data analysis pipeline architecture incorporated three types of data input; (1) sample metadata, (2) sequencing data from MinION runs, and (3) Biomeme qPCR results. Input was driven by the metadata workbook, which passed through a validation step, followed by data assimilation of sequence reads across runs, and finally the generation of pipeline-specific sample sheets. The complete pipeline architecture, from input through to Enlighten visualization output, is shown in Figure 6.

**Figure 6:**
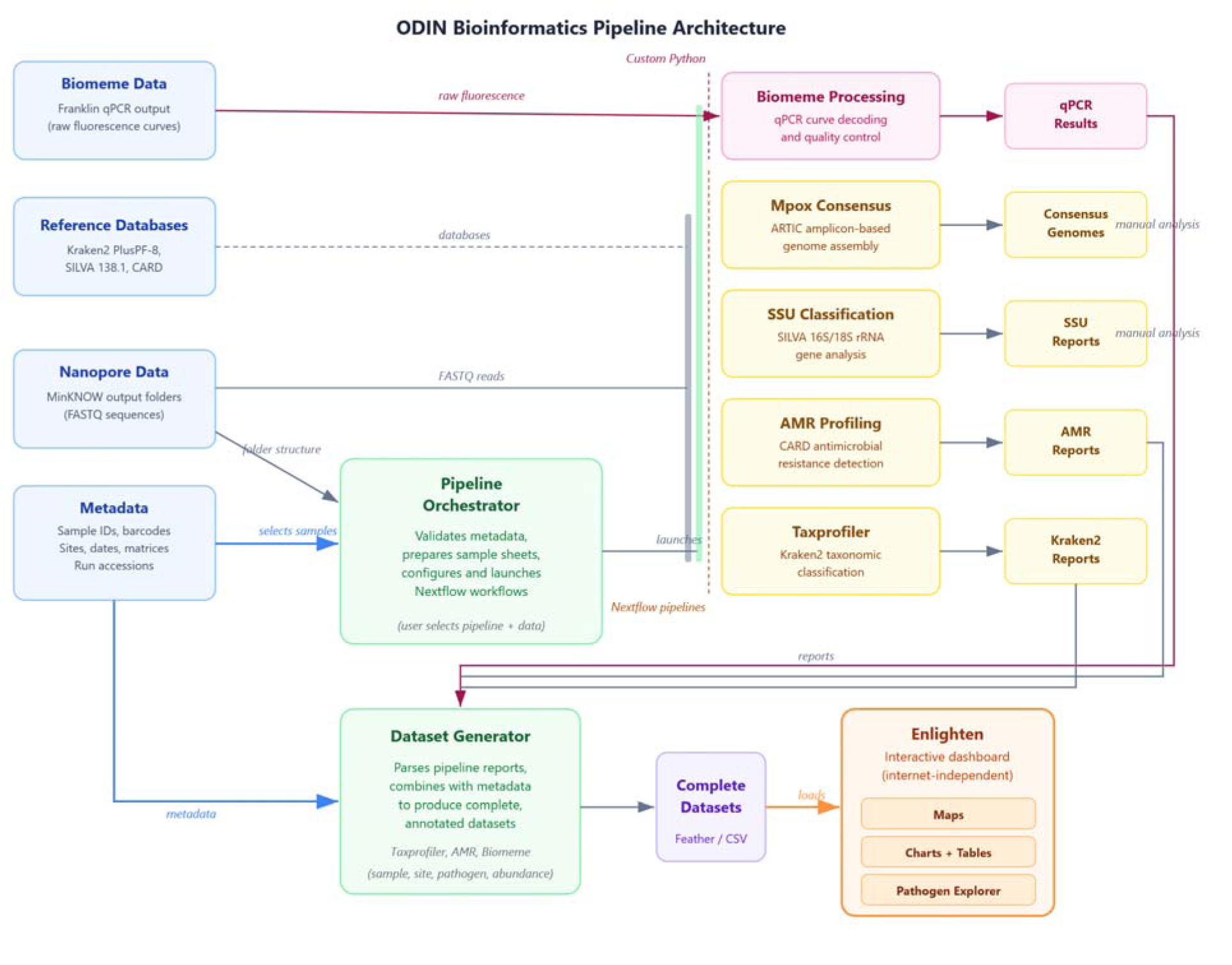
ODIN-ML Data Analysis Architecture

The data analysis architecture incorporated three bioinformatic workflows for the processing of long-read sequences from MinION sequencing runs (Table 1).

**Table 1:** Bioinformatic pipelines used during the deployment.

| Purpose | Pipeline/Tool | Description |
| --- | --- | --- |
| Preliminary screening for priority pathogens | nf-core/taxprofiler | Kraken2 taxonomic classification (Custom database), krona visualization |
| AMR detection | epi2me-labs/wf-metagenomics | AMR gene detection (CARD database) |
| Mpox characterization | artic-network/artic-mpxv-nf + squirrel | Mpox consensus calling + phylogenetic tree |

#### 2.7.1 Laptop configuration, MinKNOW/pipeline installation and ready-to-use database preparation

Laptops purchased for the ML deployment met the processing requirements to run Nanopore sequencing as well as execute bioinformatic pipelines. To execute bioinformatic pipelines, three Nextflow resource profiles were used: odin (4 CPU / 16 GB RAM / 1 h timeout) for routine samples, and odin_big (16 CPU / 31 GB RAM / 3 h timeout) for larger or more complex runs, and epi2me (Docker execution without fixed resource limits) for the AMR and Mpox pipelines.

##### MinKNOW installation

MinKNOW software (offline version: 24.11.10, subpackage versions: MinKNOW core: 6.2.8, Dorado: 7.6.8, Bream: 8.2.5, Script Configuration: 6.2.12) was provided by Nanopore and was pre-installed on ODIN laptops so that sequencing runs could be performed without internet connection.

##### Pipeline installation

All pipelines were pre-installed on ODIN laptops and tested before the deployment. Most pipelines (except for Squirrel) are containerized using Docker and orchestrated through Nextflow, running on a Windows laptop with Linux running in parallel via Windows Subsystem for Linux (WSL). In the context of this study, Windows laptops with WSL is the only configuration was used; other operating systems were not tested and may require further script adjustment before being used. This setup kept the analysis environment isolated from the host operating system and required no internet connection during analysis runs. The software stack was installed and configured prior to deployment; an initial online run pulled all required software images and pipeline assets, after which all subsequent runs operated entirely offline. Squirrel was installed via the Conda package manager.

##### Database download

For priority pathogen detection, a custom-built kraken2 database for taxprofiler was downloaded and stored locally prior to deployment. Database paths were configured via input_sheets/databases.csv. For AMR detection, the CARD database (for AMR gene annotation) and the PlusPF-8 Kraken2 database set (for taxonomic classification) were both downloaded for the epi2me-labs/wf-metagenomics workflow with the AMR detection option. For Mpox characterization, primer schemes (yale-mpox/2000/v1.0.0-cladei, yale-mpox/2000/v1.0.0-cladeii, or artic-inrb-mpox/2500/v1.0.0) for the artic-mpxv-nf workflow were downloaded from the quick-lab/primerschemes repository and stored in a local store_dir/primer-schemes/ directory. The Mpox characterization pipeline offered the following clade options: clade I, clade Ia, clade Ib, clade II, clade IIa, clade IIb.

#### 2.7.2 Preliminary screening of priority pathogens

For the screening of priority pathogens from metagenomic sequence data, the nf-core/taxprofiler bioinformatic pipeline was used ^21^. Taxonomic annotation of sequencing reads was performed using kraken2 (--run_kraken2) and a custom kraken2 database. The pipeline was run with following parameters: -profile odin, --run_kraken2, --run_krona, and a custom resource configuration (odin.config) with the “quick” option enabled for kraken2. The kraken2 database was specified via --databases flag, pointing to a deployment-specific database CSV file.

All priority pathogen results presented in this study were based on the ODIN v6 database, a custom kraken2 reference database built by NORCE team in WP4. The pathogens included in the custom database were based on the results of stakeholder surveys with partners from DRC, Tanzania, and Burkina Faso conducted as part of the ODIN project ^22^. The database was built from curated, pathogen-specific sequence collections downloaded from NCBI Datasets and NCBI RefSeq, targeting the 13 priority pathogens in the ODIN surveillance framework. The build used Kraken2 v2.1.6 (k-mer length 35, minimizer length 31, minimizer spaces 7) without a hard database-size cap, improving representation of larger eukaryotic genomes.

#### 2.7.3 Detection of AMR genes

The wf-metagenomics pipeline developed by epi2me-labs ^23^ was integrated into an orchestration script (start_nextflow_amr.sh) that handled metadata validation and pipeline invocation. This script prompted the analyst for the MinKNOW data directory, run accession, output directory, and metadata file, with previously used values offered as defaults. Metadata consistency was verified before the pipeline was launched. The fastq_pass directory from the selected run accession was passed directly to the pipeline as input. The pipeline was run with the following parameters: --amr --amr_db card --database_set PlusPF-8.

#### 2.7.4 Mpox characterization

Sequencing data were analyzed using two publicly available bioinformatic tools: the artic-mpxv-nf workflow for generating consensus MPXV genomes from pooled tiled amplicons, and Squirrel for sequence alignment and maximum-likelihood phylogenetic inference ^24–33^. The artic-mpxv-nf and Squirrel tools were integrated into a single orchestration script (start_mpox.sh) that guided the analyst through an interactive selection menu for Mpox clade and primer scheme version, with fixed defaults pre-selected (cladeii and artic-inrb-mpox/2500/v1.0.0 respectively). Prior to pipeline execution, a sample sheet was generated by create_mpox_samples_csv.py, which matched barcode directories in the fastq_pass folder against the metadata workbook for the selected run accession and produced a CSV file with three columns: barcode, sample alias, and sample type (test_sample, positive_control, negative_control, or no_template_control). Barcodes present in the sequencing data but absent from the metadata were skipped and reported. This sample sheet was passed to the Nextflow pipeline via the --sample_sheet parameter alongside --fastq, --clade, --scheme_version, and --store_dir. Following Nextflow execution, the script automatically activated the Squirrel conda environment and ran phylogenetic inference with the flags --run-apobec3-phylo --include-background. This design allowed analysts without command-line experience to run the full consensus-to-phylogeny workflow by responding to a small number of guided prompts.

#### 2.7.5 Biomeme qPCR data processing

A custom Python pipeline was developed to process Cq values exported from the Biomeme Franklin instrument. The pipeline extracted Cq values from instrument output files, merged them with metadata and assay-specific parameters derived from standard curves (slope M, intercept B, efficiency E, threshold T), and calculated gene copy abundance estimates for each target ^19^.

Each qPCR measurement was assigned a quality flag indicating data reliability: flag 1 (Good — both qPCR replicates exhibited non-zero replicate Cq values), flag 2 (Questionable — one out of two qPCR replicates exhibited a non-zero Cq value), flag 3 (Negative — no amplification), or flag 4 (NTC contamination — A non-zero Cq value was observed in one of the no-template control reactons). A derived presence column classifies each measurement as true (flag 1 + abundance > 0), true_x (flag 2 + abundance > 0, i.e. tentative detection), or false (all other cases). This framework enables transparent downstream filtering by detection confidence.

#### 2.7.6 Visualization of the constructed datasets

Enlighten is a Docker-based web visualisation platform consisting of a REST API service and a web frontend, served locally on the ODIN laptops and requiring no internet connection. The platform was used to load structured datasets generated by postprocessing modules for pathogen profiling and qPCR data. Taxonomic profiling datasets included fields for sample name, barcode, taxon, read counts, relative abundance, and a binary pathogen priority flag derived from a curated pathogen database. The dashboard presented interactive bar charts (by sampling site, priority pathogen group, genus, and target group), an interactive map, and a tabular data view. For Biomeme qPCR results, the dashboard connected Cq values to corresponding metadata entries, applied the quality flag filtering described in §2.4, and displayed gene copy estimates on interactive maps and barplots for spatial and temporal trend visualization.

### 2.8 Training component

#### 2.8.1 Overview of participants during the training

Training on ML modules was conducted in parallel with sample analysis and much of the data presented from the ML deployment was generated by African partners participating in training. A team of seven NORCE researchers (four on-site and three providing remote support) conducted training and provided troubleshooting support throughout the month-long mobile laboratory (ML) deployment. In addition, one researcher from each of the University of Helsinki (UH) and the Finnish Institute for Health and Welfare (THL) helped with training during the deployment. Over the course of the 4-week deployment, training sessions were conducted with a total of 25 researchers: 13 from Tanzania, 6 from Burkina Faso, 5 from DRC, and 1 from Ethiopia. Approximately 5–8 African researchers visited the ML each week, receiving hands-on training on each module. The ML lab was also visited by Public Health Authorities in Tanzania during training sessions to discuss the scope of ML activities and opportunities for expansion.

#### 2.8.2 Lab workflows and bioinformatic pipelines

##### Training modules covered

During the ML deployment, training modules included: sample collection and filtration, NA extraction, Biomeme qPCR, metagenomic sequencing, Mpox tiling amplicon sequencing. The modular design of the mobile laboratory directly supported the training program. For example, participants with different specializations could be trained on individual modules without requiring a comprehensive overview of the entire laboratory workflow. This modular design enabled parallel training sessions to be conducted, making the training more efficient. The training also included pipeline installation, executing offline bioinformatic scripts, and Enlighten dashboard operation. Trainees had the opportunity to analyze data, exchange information and discuss results onsite. A proportion of participants from DRC and Burkina Faso were French speaking. To accommodate French-speaking researchers, training protocols were translated using AI-assisted translation tools, with accuracy confirmed by a French-speaking team member from Norway. This approach, combined with hands-on demonstration, successfully overcame the language barrier.

#### 2.8.3 Orchestration scripts for users with limited command-line experience

The orchestration scripts were designed to minimise the risk of user error during field operation, where analysts may have limited command-line experience. Several safeguards and usability features were incorporated (https://github.com/ODIN-consortium):

- **Persistent configuration**: All file paths and settings entered by the analyst are automatically saved to a user-specific configuration file (∼/.odin_ml/odin_paths.txt) at the end of each run and reloaded as defaults at the start of the next, eliminating the need to re-enter paths repeatedly.
- **Fallback defaults**: If no user configuration exists, scripts fall back to a set of deployment-specific defaults shipped with the pipeline, ensuring the system is immediately runnable after installation without any manual configuration.
- **Path validation with re-prompting**: All path inputs are validated before any pipeline step is executed. If a specified directory or file does not exist, the script prompts the analyst to re-enter the path rather than proceeding with an invalid argument, preventing failures deep into a multi-hour pipeline run.
- **Windows/WSL path compatibility**: Paths copied from Windows Explorer (e.g. C:\data\run1) are automatically converted to their WSL equivalents (/mnt/c/data/run1), removing a common source of confusion for analysts working across both Windows and Linux environments.
- **Output directory protection**: Scripts check whether the designated output directory already contains data before starting. If it is non-empty, execution is halted with a clear error message, preventing accidental overwriting of existing results.
- **Interactive run accession selection**: Rather than requiring analysts to type the full run accession path, scripts scan the MinKNOW data directory and present a numbered menu of valid sequencing runs, from which the analyst selects by number.
- **Metadata pre-validation**: Metadata consistency is verified by verify_nanopore_metadata.py before any Nextflow pipeline is launched. If validation fails, the script exits immediately with a descriptive error, avoiding wasted compute time on incorrectly configured runs.
- **Guided parameter selection**: For pipelines with configurable biological parameters, such as Mpox clade and primer scheme, analysts are presented with a numbered menu of supported options with a pre-selected default, so that the most common choice requires only pressing Enter.
- **Color-coded console output**: Error messages are displayed in red, successful completion messages in green, and warnings in yellow, making it straightforward to identify the outcome of each pipeline step at a glance.

#### 2.8.4 Post-deployment follow-up

Certificates of completion were issued to all 25 African participants after the deployment and post-deployment surveys were sent out to gather feedback from these participants.

#### 2.8.5 Analysis of survey data analysis on trainees’ perception on ML in ODIN

An analysis was conducted using responses from stakeholders who reported direct participation in ODIN ML deployment and training activities to investigate perspectives on the pilot implementation of the mobile wastewater surveillance lab and identify areas for improvement. Agreement was calculated as the percentage of respondents selecting a given response option among all valid responses to the corresponding question. Questions permitting multiple selections, including use cases, preferred outputs, and implementation requirements, as well as respondent profile questions and open-ended responses, were excluded from this analysis. Data processing, aggregation, summarisation, and visualisation were performed in the R statistical computing environment using the packages readxl, dplyr, janitor, ggplot2, stringr, and patchwork.

## 3 Results

### 3.1 Preliminary screening for priority pathogens

Taxonomic profiling with nf-core/taxprofiler (Kraken2, ODIN v6 reference database) was applied to 83 environmental samples collected in Tanzania, Burkina Faso, and DRC between January and September 2025. Results are summarized with annotated heatmaps for each country, where cells report detected/total samples and colour intensity depicts mean reads per million classified reads (RPM, log10 scale) for positive detections. It is important to note that these results are based on preliminary screening using curated priority pathogen databases and require further confirmation tests.

In Tanzania samples (n = 26), priority bacteria in the potentially AMR-associated group were detected at high prevalence across all five sample types (drinking water, surface water, wastewater and soil; Figure 7). *Salmonella* spp. and *Enterobacter* spp. were the most abundant taxa detected, present in all 26 samples with mean abundances of 178,512 and 126,713 RPM, respectively. *Acinetobacter baumannii* and *Pseudomonas aeruginosa* were similarly widespread (detected in 36/38 and 37/38 samples), consistent with environmental circulation of opportunistic AMR-associated organisms. Among other human pathogens, *Vibrio cholerae* was detected in 37/38 samples and *Cryptosporidium spp*. in 37/38 samples including all 13 river water samples and all 11 drinking water samples, indicating high environmental prevalence across the sampled sites. *Giardia* spp. was detected in 36/38 samples.

**Figure 7:**
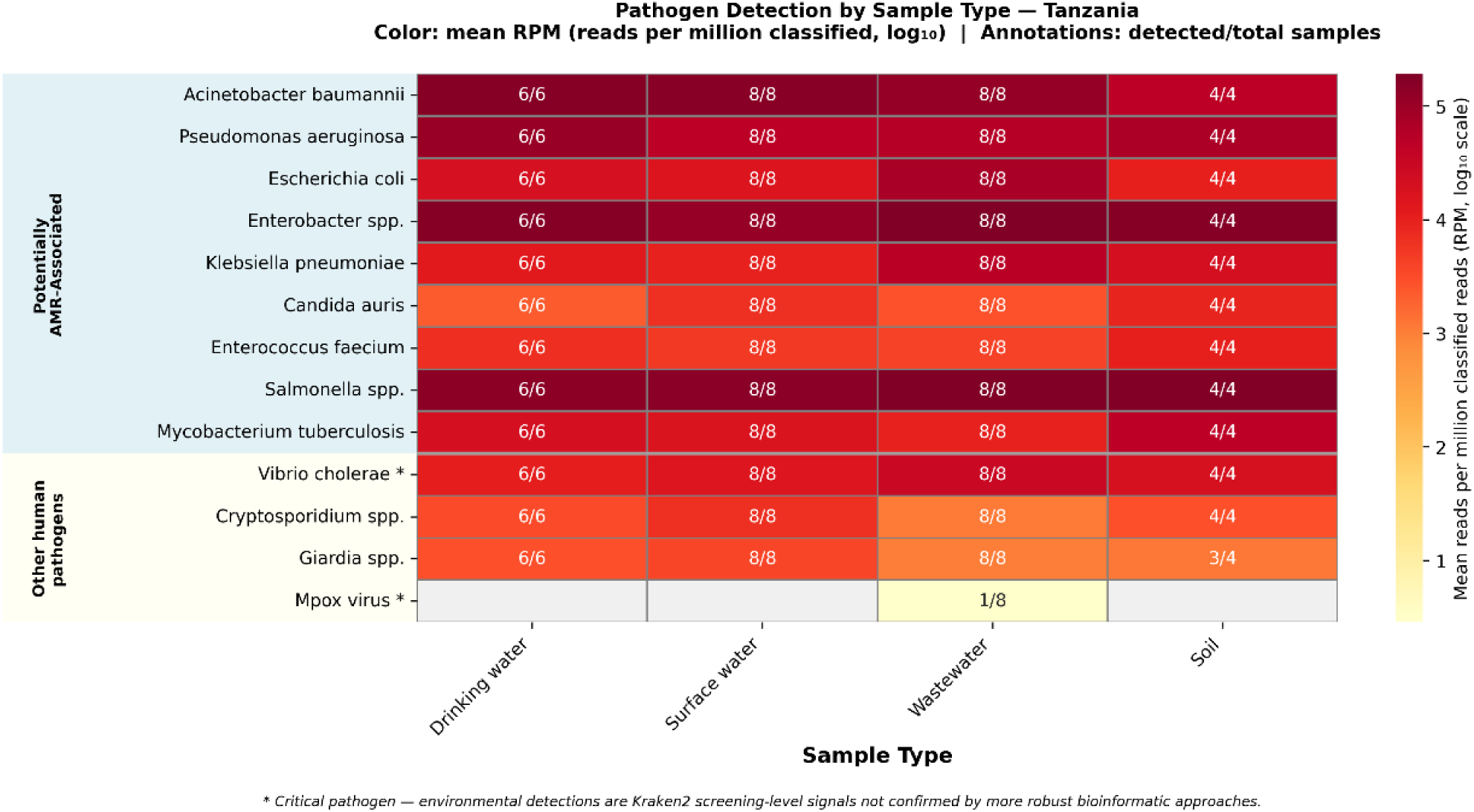
Priority pathogen detection by sample type — Tanzania (26 samples, January–September 2025). Cell annotations show detected/total samples. Colour scale: mean RPM (log₁₀). Grey cells = no detections. Of note, these results are preliminary screening level which require further testing to confirm their presence, especially for critical pathogens marked with an asterisk (*).

Burkina Faso samples (n = 21) included wastewater, surface water, and drinking water (Figure 8), potentially AMR-associated bacteria showed similarly high detection rates to those observed in Tanzania samples. *Salmonella spp.* had the highest mean abundance (238,709 RPM), followed by *Enterobacter* spp. (162,298 RPM) and *Pseudomonas aeruginosa* (121,150 RPM). *Candida auris*, an emerging multidrug-resistant fungal pathogen, was detected in 15/21 samples, though at substantially lower abundance (mean 1,291 RPM) than the bacterial AMR taxa. *Vibrio cholerae* was detected in 19/21 samples across all sample types.

**Figure 8:**
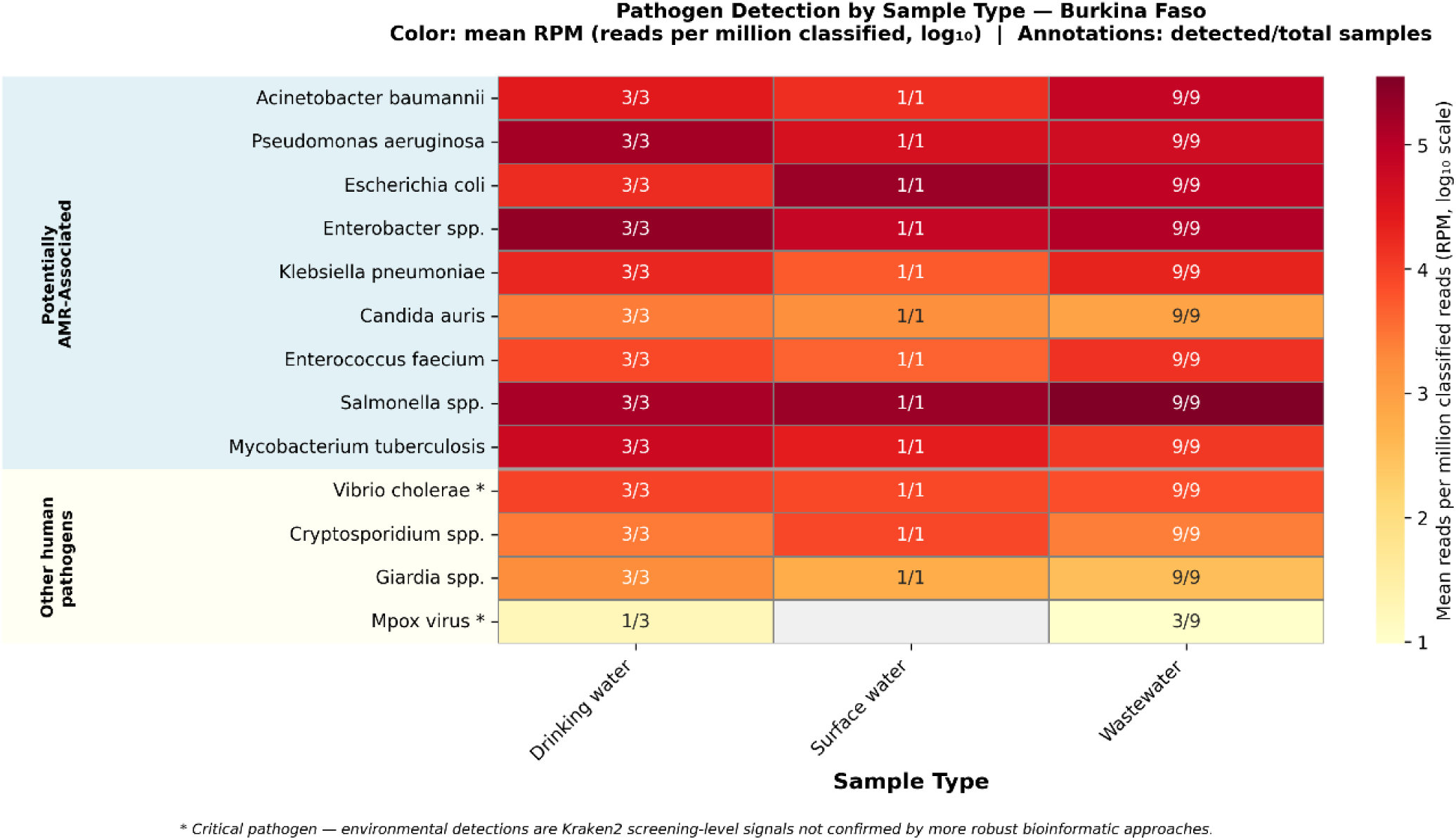
Priority pathogen detection by sample type — Burkina Faso (13 samples, January–September 2025). Cell annotations show detected/total samples. Colour scale: mean RPM (log₁₀). Grey cells = no detections. Of note, these results are preliminary screening level which require further testing to confirm their presence, especially for critical pathogens marked with an asterisk (*).

In DRC environmental samples (n = 14; **Figure 9**), among potentially AMR-associated pathogen group, *Escherichia coli* and *Salmonella* spp. were the most abundant (mean RPM 188,850 and 179,267, respectively), each detected in 13–14 of 14 samples. *Mycobacterium tuberculosis* was detected in 11/14 samples across all sample types, consistent with the high TB burden in the region. *Vibrio cholerae* was detected in 9/14 samples including both surface water samples, consistent with documented cholera transmission in eastern DRC. Detection rates for *Candida auris* and *Enterococcus faecium* were lower than in the other countries (4/14 and 5/14, respectively).

**Figure 9:**
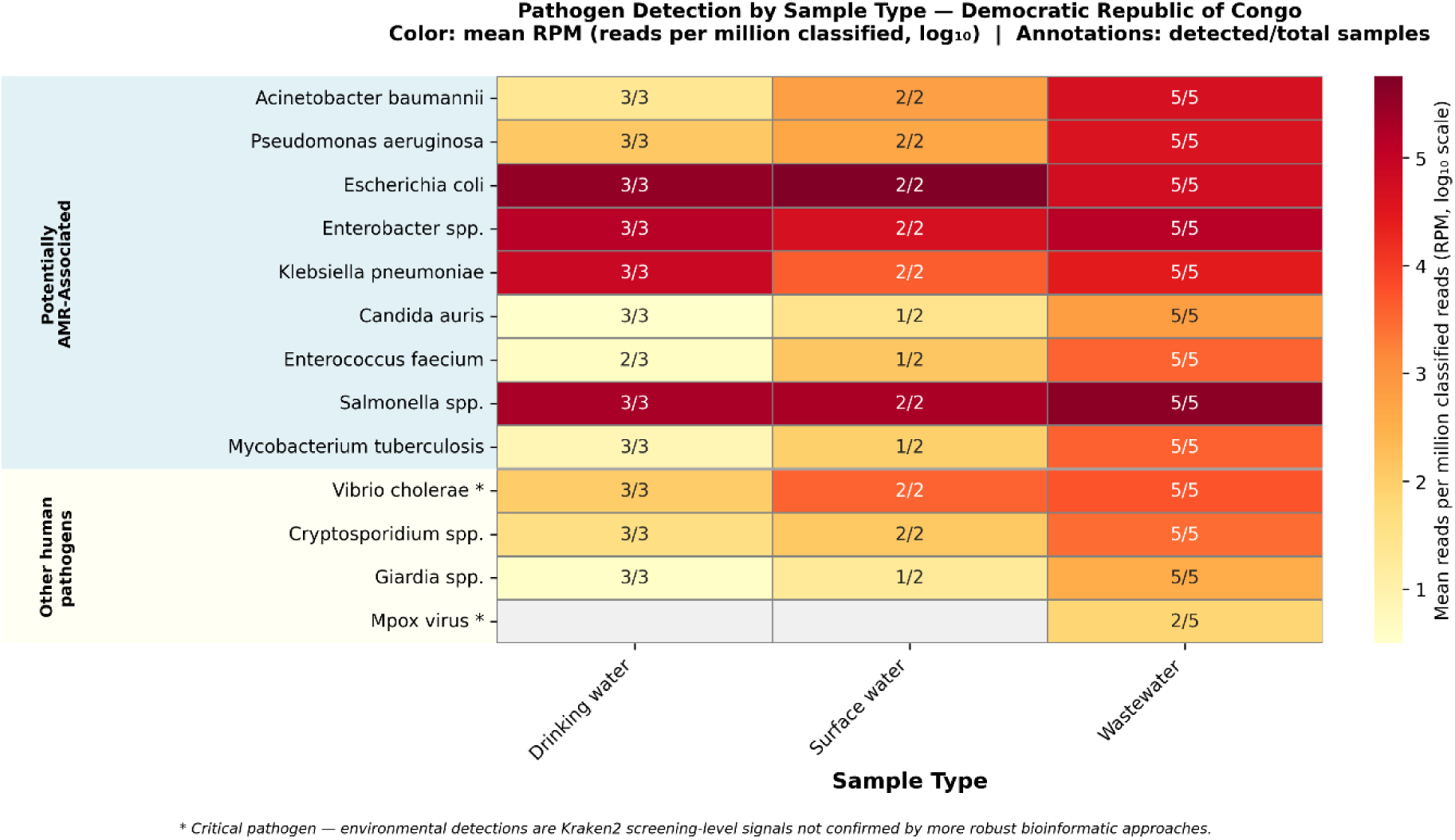
Priority pathogen detection by sample type — Democratic Republic of Congo (10 samples, January–September 2025). Cell annotations show detected/total samples. Colour scale: mean RPM (log₁₀). Grey cells = no detections. Of note, these results are preliminary screening level which require further testing to confirm their presence, especially for critical pathogens marked with an asterisk (*).

### 3.2 AMR profile analysis

The presence and abundance of antimicrobial resistance (AMR) genes was assessed using metagenomic AMR profiling against the CARD database. A total of 50 samples were analysed, including environmental samples from Tanzania (n = 22), Burkina Faso (n = 17), and DRC (n = 11). The 50 samples analyzed for AMR genes represented a subset of the 101 samples collected across the three countries (Table S1). Samples were included in AMR profiling only if they were sequenced under Protocol P2 (the standardised metagenomic workflow) and clinical samples were excluded from this analysis, ensuring comparability of AMR gene quantification across sites. This yielded 65 unique AMR resistance profiles. The top 25 profiles by mean read count are shown in the clustered heatmap (Figure 10), with sample types grouped by country and values expressed as log10 (mean reads + 1).

**Figure 10:**
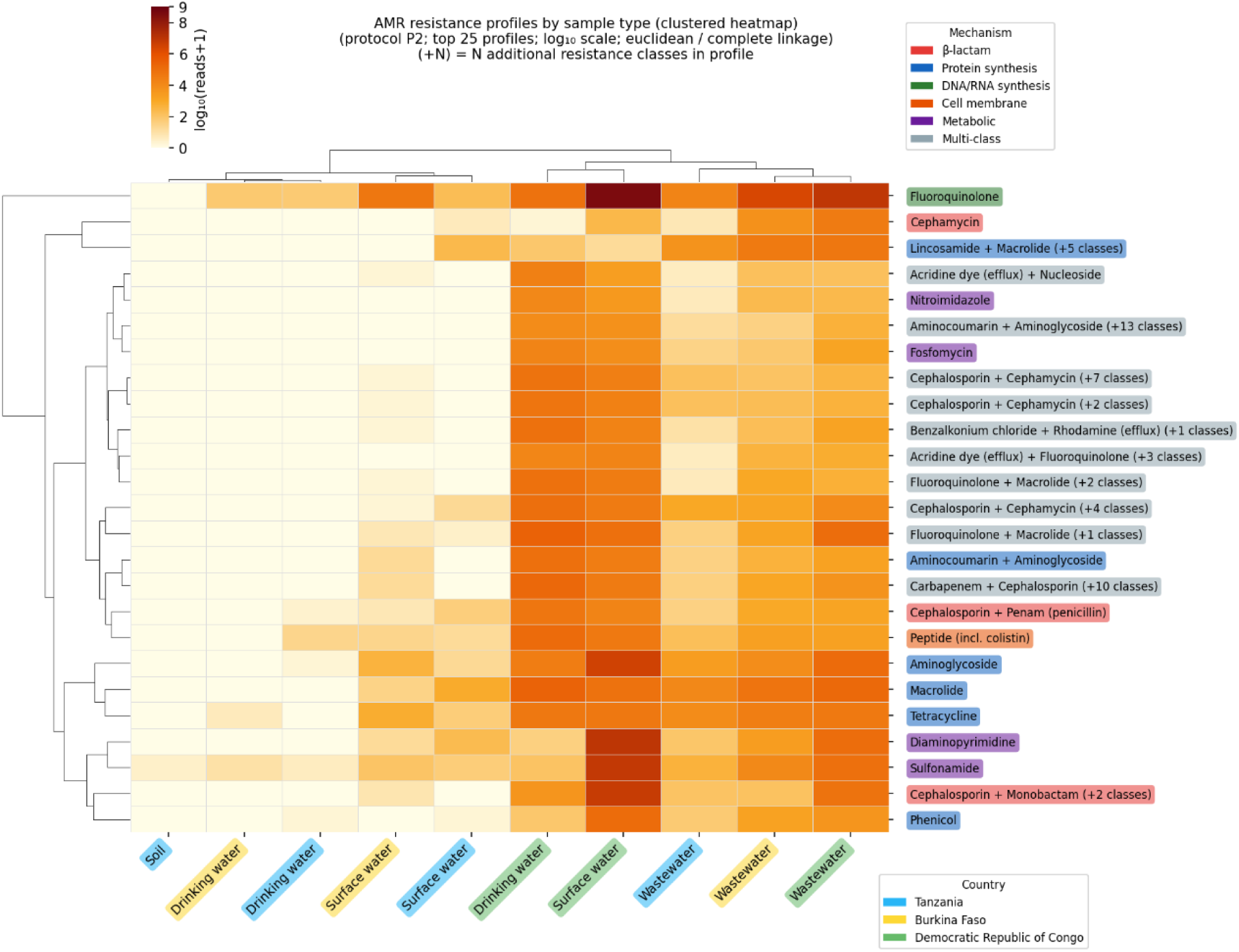
AMR resistance profiles by sample type (top 25 profiles; 50 samples across Tanzania, Burkina Faso, and DRC; January–September 2025). Rows: resistance profile (primary drug class codes; see legend). Columns: country–sample type combinations, clustered by Euclidean distance / complete linkage. Colour: log₁₀(mean reads + 1). BF = Burkina Faso, DC = DRC, TZ = Tanzania.

Fluoroquinolone resistance genes were the most dominant AMR signal observed across all countries and sample types, with the highest read counts in Burkina Faso and DRC wastewater, and consistent detection spanning drinking water, surface water, and wastewater. AMR genes for fluoroquinolone were detected in samples from all three countries across every sample type tested, suggesting ubiquitous environmental dissemination of quinolone resistance in the investigated regions. Complex multi-drug resistance (MDR) profiles combining AMR genes for fluoroquinolones with beta-lactams, aminoglycosides, phenicols, and other drug classes were also prevalent, particularly in Burkina Faso and DRC wastewater samples, reflecting the diverse AMR gene pools in wastewater-impacted environments.

Multi-drug resistance profiles (genes conferring resistance to ≥3 drug classes) were detected in 16/22 Tanzania samples (73%), 14/17 Burkina Faso samples (82%), and all 11 DRC samples (100%). In DRC samples, the high MDR prevalence is consistent with the challenging healthcare context and limited antibiotic stewardship resources. The aminoglycoside; aminocoumarin profile (amnc;amng) was notably enriched in Burkina Faso wastewater, while peptide resistance (pptd, including colistin-associated ICR-Mo) was detected across all three countries, raising concern given the use of colistin as a last-resort antibiotic. Sulfonamide, diaminopyrimidine, and aminoglycoside resistance were each among the most abundant single-class profiles by read count, consistent with heavy historical use of these drug classes in sub-Saharan Africa.

Hierarchical clustering of sample types identified a high-intensity cluster comprising Burkina Faso and DRC wastewater samples, DRC drinking water, and DRC surface water, and a lower-intensity cluster containing the remaining Tanzania and Burkina Faso sample types. This separation suggests substantially higher AMR signal in DRC-associated matrices in this dataset, warranting confirmation with larger sample sets.

### 3.3 qPCR analysis of priority pathogens

qPCR screening on the Biomeme Franklin platform detected *E. coli*, *K. pneumoniae*, *V. cholerae,* and MPXV targets in a subset of field samples; whereas no positives were observed for *S. typhi*, *Orthopox spp*, *MPXV clade I*, or *MPXV clade Ib* qPCR assays (Table S1). Of the 59 samples tested for *K. pneumoniae*, 45 were positive. For *V. cholerae*, 4 out of 59 tested samples were positive. Only three samples were tested for *E. coli* using qPCR, of which one sample was positive. Of the 15 samples tested with the generic *MPXV* qPCR assay, two were positive; however, *MPXV* detections were observed exclusively in clinical samples from the DRC, and no environmental samples were confirmed to be qPCR positive during Biomeme qPCR testing (Table S1).

Compared with metagenomic profiling, qPCR coverage was sparser across sites and sample types, but overlapping samples showed good qualitative agreement for detections of *K. pneumoniae*, *V. cholerae,* and *E. coli*. The clinical samples from the DRC were not subjected to metagenomic sequencing; therefore, we could not compare qPCR and metagenomic results for *MPXV* (Table S1).

### 3.4 Mpox detection

Preliminary MPXV signals were detected in a subset of environmental samples during Kraken2-based metagenomic screening. However, these findings could not be confirmed by downstream read mapping, BLAST analysis, qPCR, or MPXV tiling amplicon sequencing. Taken together, the results provide no convincing evidence for the presence of MPXV in the investigated environmental samples.

During the validation of our Mpox tiling amplicon methods, we were able to precisely characterize the Mpox clade II strain from Slovenia in spiked samples as previously described ^19^. Subsequently, this approach was validated with other Mpox clades (clade Ia, clade Ib and clade IIb) obtained from WHO BioHub (https://www.who.int/initiatives/who-biohub). The protocol was also able to correctly identify each Mpox clade when spiked into WW samples. While clade Ib and IIb were accurately projected on to the phylogenetic tree after one round of PCR amplification, clade Ia was more difficult and required two rounds of PCR amplification for accurate identification. In addition to inactivated viruses, this tiling amplicon sequencing approach was also performed on pre-extracted DNA of Mpox clade II obtained from a Dutch distributor, which also required two rounds of PCR amplification before accurate projection on the phylogenetic tree (supplemental document 1).

During the deployment, tiling amplicon sequencing was performed on both environmental and clinical samples. However, environmental samples did not yield any high-quality consensus sequences for subsequent construction of a phylogenetic tree. Only consensus sequences from clinical samples were projected on the phylogentic tree, which were clustered well and more closely related to reference Mpox genomes within clade Ia than clade II (**Figure 11**). Two samples (DCTPHP1 & DCTPHP3) were successfully mapped on to the phylogenetic tree after one round of PCR amplification, while two additional samples (DCTPHP4 & DCTPHP5) required two rounds of PCR amplification before they could be phylogenetically mapped. DCTPHP2 was still missing from the tree after two rounds of PCR amplification. These results were consistent with data provided by the DRC team indicating that these samples had been independently identified as clade Ia infections. The qPCR assays using generic primers (G2R) were performed on three clinical samples and generally agreed with the above amplicon tiling approach: DCTPHP1 & 3 were strongly positive using the generic Mpox assay; although no positives were observed using clade-specific qPCR assays among the clinical samples that were tested.

**Figure 11:**
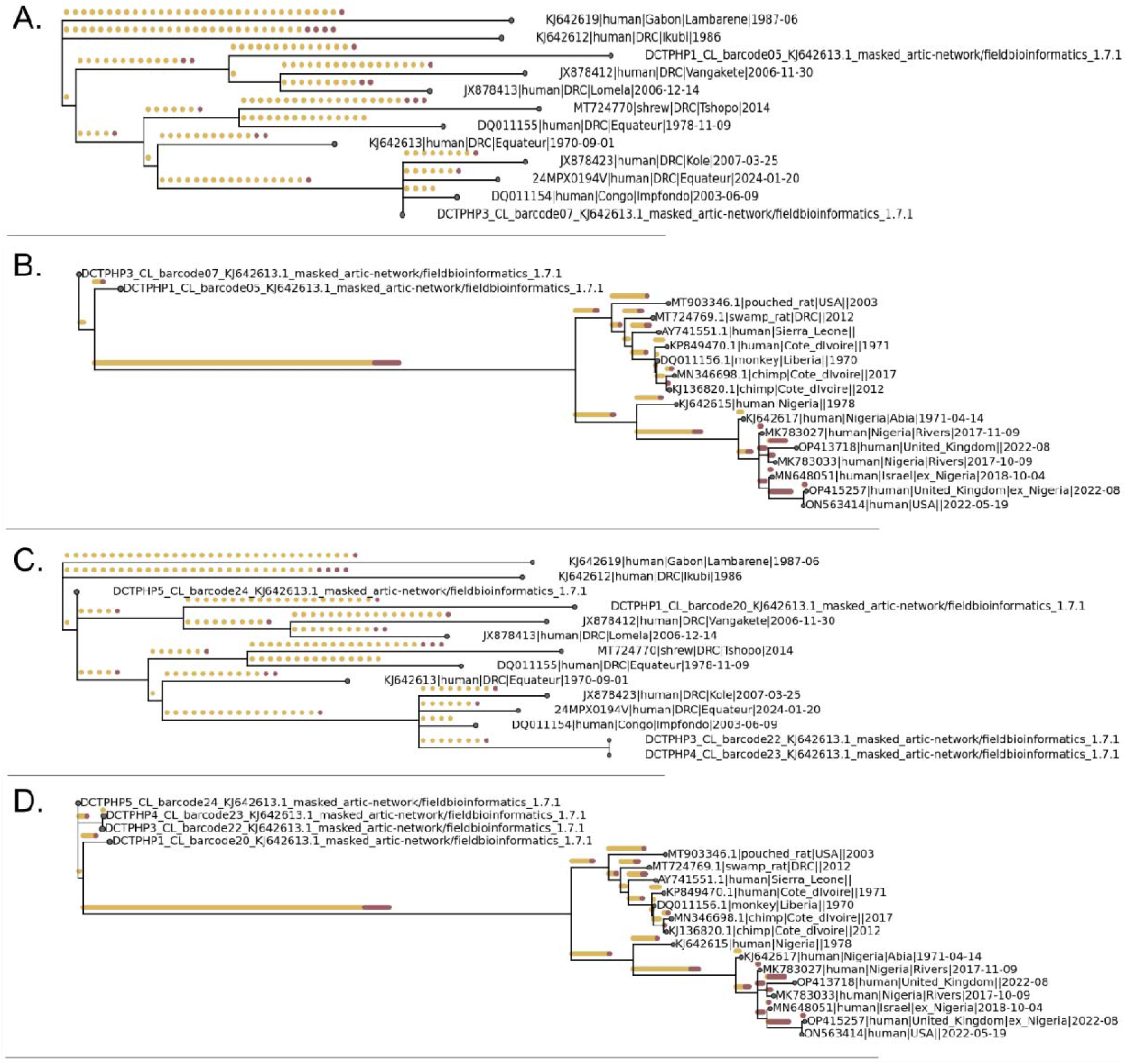
Phylogenetic alignment of Mpox DNA from confirmed clinical cases in DRC with reference sequences from clade Ia (A and C) or clade II (B and D). Two samples (DCTPHP1 & 3) were successfully shown on the phylogenetic tree after one-round PCR amplification (A and B), while two additional samples (DCTPHP4 & 5) were added to the tree after two-round PCR amplification (C and D).

### 3.5 Performance of automated data analysis and visualization workflow

Enlighten provided standalone, on-site integration and visualization of results from all bioinformatic pipeline outputs and Biomeme qPCR data.

During the deployment, Enlighten visualization templates were updated remotely by the WP4 team at NORCE (Norway) and pushed to the field laptop as new requirements emerged from real-world use. This included adjustments to how pathogen groups were categorized and displayed, new chart configurations, and modified data table layouts — all delivered without requiring the field team to modify any configuration files. The ability to evolve the visualization layer during an active deployment illustrates the flexibility of the template-driven approach.

Feedback from training participants and visiting government health officials indicated that the interactive visualizations — particularly geographic distribution maps and priority pathogen bar charts — were considered directly useful for public health decision-making, more so than raw tabular output or classification reports alone.

The Enlighten data visualization platform was deployed on the analysis laptop throughout the field campaign, providing real-time, internet-independent integration of results from all four pipeline outputs and the Biomeme qPCR instrument. Datasets from each sequencing run were automatically ingested as Nextflow pipelines completed, making updated results available within minutes of pipeline execution. Two representative dashboard views are shown in Figures 12 and 13.

**Figure 12:**
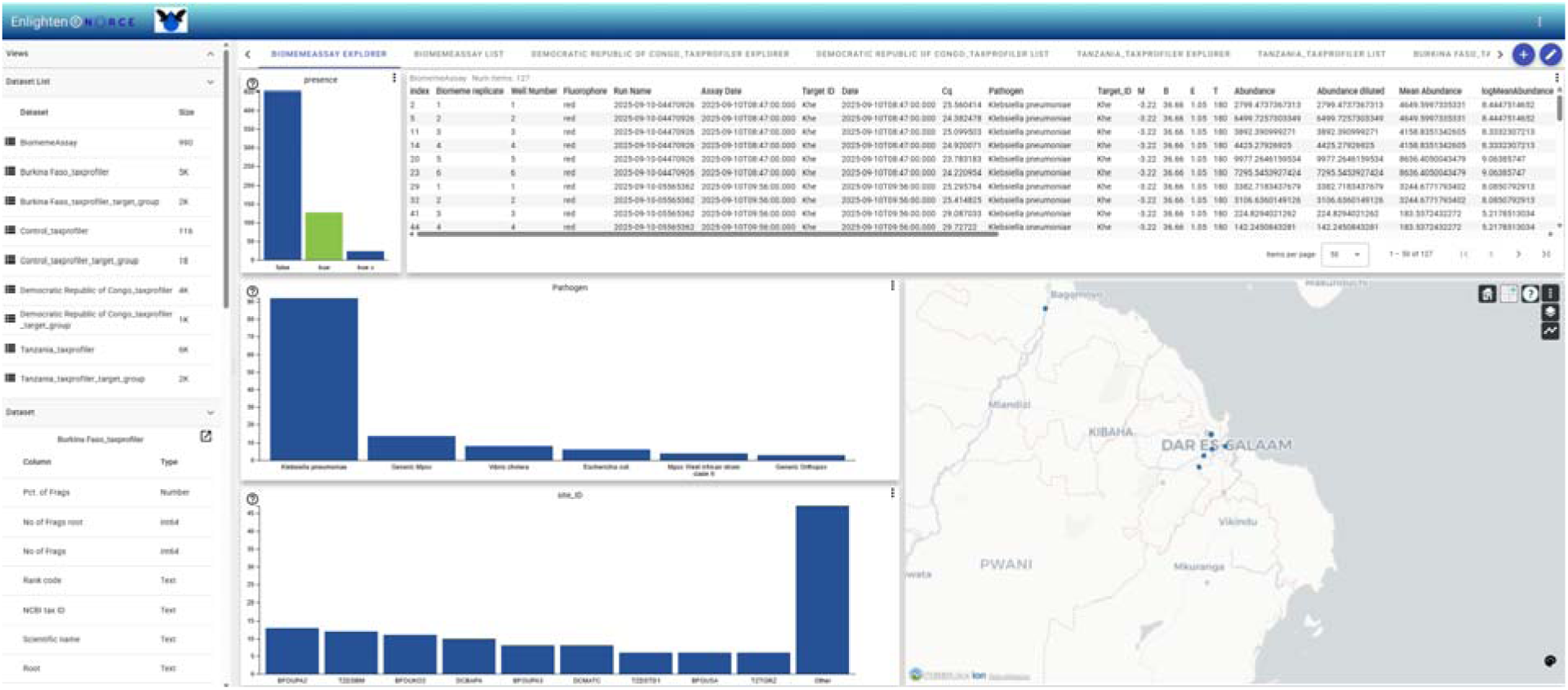
Enlighten Biomeme Assay Explorer — integrated view of Biomeme qPCR results showing the presence/absence chart (top left), tabular assay data with Cq values and pathogen identifications (top right), pathogen frequency bar chart (centre left), site frequency chart (centre right), and geographic map of sampling sites (right). Screenshot from Tanzania deployment, September 2025.

**Figure 13:**
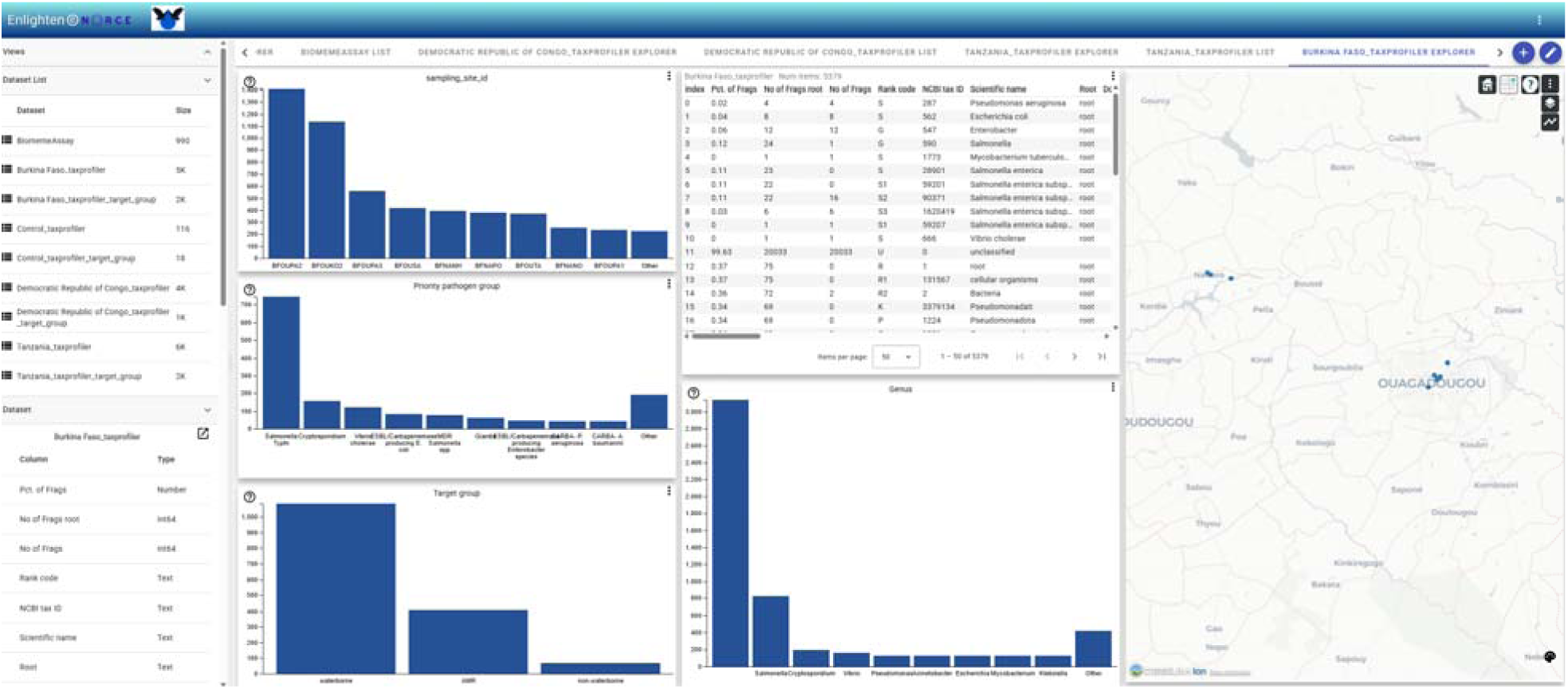
Enlighten Taxprofiler Explorer — integrated view of metagenomic classification results for the Burkina Faso dataset, showing sampling site read counts (top left), priority pathogen group detection frequencies (centre left), target group summary (bottom left), raw Kraken2 classification table (top right), genus-level bar chart (centre right), and geographic map (far right). Screenshot from Burkina Faso dataset.

The Biomeme Assay Explorer (Figure 12) provided a consolidated view of qPCR results across all Biomeme Franklin runs, integrating assay data, Cq values, sample metadata, and geographic coordinates. Presence/absence bar charts, a searchable data table, and an interactive map enabled field operators to rapidly assess detection patterns across sample types and sites without requiring data export or manual post-processing.

The Taxprofiler Explorer (Figure 13) displayed metagenomic classification results at the read level, providing priority pathogen group counts, target group summaries, genus-level breakdowns, and the raw Kraken2 classification table. Colour-coded site maps enabled visual cross-referencing of detection patterns with sampling geography. The dashboard supported on-site interpretation of results by field scientists without requiring command-line access to pipeline outputs.

### 3.6 Trainees’ feedback on the mobile surveillance laboratory in ODIN and future implementation

The feedback outlined here represents only a subset of a broader stakeholder questionnaire, the full results of which will be presented in a comprehensive report currently in preparation. Responses from participants involved in training (n=12) highlighted the significant potential of the ODIN mobile laboratory platform to strengthen environmental and public health surveillance. Trainees consistently emphasized the benefits of rapid, field-based diagnostics, near real-time analysis, and the ability to generate actionable data directly at the point of need. The platform was perceived as particularly valuable for early outbreak detection, antimicrobial resistance monitoring, environmental surveillance, and emergency response activities in remote and underserved areas. Several respondents noted that the mobile laboratory approach could reduce delays associated with sample transportation and centralized testing, allowing faster identification of public health threats and more timely interventions. Feedback from trainees further indicated that the platform has strong potential to support evidence-based decision-making. Participants described how rapid access to diagnostic and genomic data could help authorities identify transmission hotspots, prioritize water treatment measures, strengthen infection prevention and control activities, target outbreak responses, and improve preparedness for future health emergencies. Many respondents also highlighted the relevance of the One Health approach and the integration of environmental, human, and antimicrobial resistance surveillance within a single operational framework. Regarding future implementation, respondents expressed strong support for wider adoption of the platform while emphasizing the need for sustainable funding mechanisms, continued training and capacity building, integration with national surveillance systems, and adequate infrastructure and logistical support. The importance of developing local expertise, ensuring regulatory acceptance, and establishing long-term operational and maintenance plans was repeatedly highlighted. Several trainees suggested that mobile laboratory units could be strategically deployed in outbreak-prone areas, border regions, ports of entry, and areas with limited access to conventional laboratory facilities to support both routine surveillance and emergency response activities. Overall, trainees viewed the ODIN mobile laboratory platform as a promising and innovative tool capable of enhancing outbreak preparedness, improving surveillance coverage, accelerating public health responses, and supporting more effective disease prevention and control efforts, provided that key implementation and sustainability requirements are addressed. The outcomes from the survey and responses to selected closed-ended single-choice questions addressing environmental surveillance familiarity (Q7), relevance for public health surveillance (Q11), decision support (Q15), timeliness improvement (Q16), decision impact (Q17), output interpretation (Q19), policy suitability (Q21), and implementation feasibility (Q22) are presented in the figure 14.

**Figure 14:**
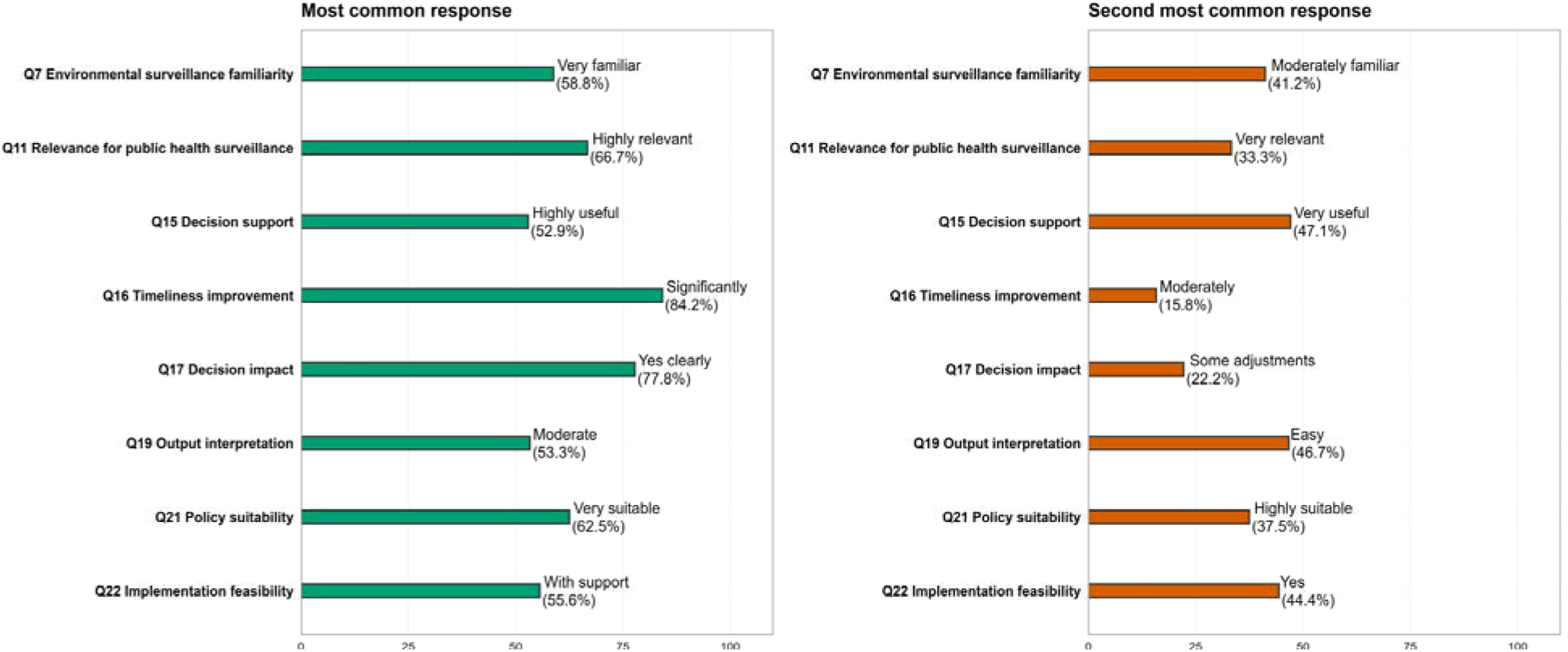
Agreement with the two most frequently selected response categories among deployment and training participants. The analysis includes all closed-ended single-choice questions considered relevant for assessing perceptions of the ODIN mobile laboratory approach (Q7, Q11, Q15, Q16, Q17, Q19, Q21 and Q22). Multi-response questions, respondent demographic questions, and qualitative free-text responses were excluded

## 4 Discussion

Wastewater and broader environmental surveillance provide a population-level fingerprint of community health. Between 1970 and 2024, wastewater surveillance efforts identified 107 distinct pathogenic species ^34^. Numerous studies have emphasized the role of wastewater in monitoring human pathogens and screening for AMR ^34–38^. While wastewater plays a central role in the fight against infectious diseases, other environmental matrices provide additional dimensions and allow higher resolution to the disease emergence landscape ^39–41^. For this reason, our study was expanded to include a broad range of samples: wastewater, surface water, drinking water and soil. This comprehensive design enabled not only the detection of a broad spectrum of clinically relevant, emerging, and AMR-associated pathogens but also the ability to track them before they reach the healthcare systems.

The untargeted metagenomic sequencing approach has considerable potential for the simultaneous detection of target pathogens. Our preliminary screening suggests that all priority pathogens, except for Mpox, were consistently detected across all sample types in the three countries. In general, these priority pathogens were found with highest abundance in wastewater and soil, followed by surface water and finally in drinking water. This pattern is consistent with concentrated human fecal inputs in wastewater and mixed-source contamination in wastewater-linked environments ^42^. Drinking water showed lower priority pathogen diversity overall, but key indicators including *Salmonella* spp. and *Vibrio cholerae* were still detected in several instances. It is important to note that these observations were results of our preliminary screening, which were subjected to further confirmation testing. Metagenomic sequencing with Oxford Nanopore Technology generated longer reads than metabarcoding, providing higher species-level resolution, reducing biases and enabling detection of multiple pathogen classes simultaneously ^43,44^. The kraken2-based taxonomic classification against the custom ODIN database, which includes a curated set of priority pathogen genomes tailored to the regional context, was a key determinant of detection sensitivity. Low-abundance reads were prone to be misclassified as seen in Mpox. Although this study was not designed to define analytical detection thresholds, our findings emphasize the importance of establishing validated criteria for interpreting low-abundance metagenomic signals. Future work should focus on determining read-count thresholds that balance sensitivity and specificity and on defining when orthogonal confirmation is required for low-level detections. Cross-validation against Biomeme qPCR results, where both methods independently detected the same pathogen, substantially increased confidence in positive calls.

As Mpox outbreak was declared in DRC in April 2024, an emergency fund was allocated to advance genomic surveillance and wastewater-based epidemiology for Mpox (https://cordis.europa.eu/project/id/101195186). In this study, multiple approaches were used for Mpox detection including metagenomic (taxprofiler), targeted (Biomeme qPCR) and tiling amplicon sequencing approaches (artic + squirrel). A small number of reads in some samples were mis-classified as Mpox by kraken2 tool (this tool is part of taxprofiler pipeline). Kraken2 tool is widely used for taxonomic classification because it is fast, efficient and optimized for large-scale metagenomic data ^45,46^. However, this speed and efficiency trade-off make Kraken2 more susceptible to false positives and classification errors. Misclassification occurs when queried reads are mapped to the wrong taxon, commonly because their true source genome is absent from the reference genome or these sequences are found in conserved DNA regions ^47^. This approach is more problematic for long-read sequencing technologies when long-read sequences used with k-mer-based taxonomic classifiers can result in apparently confident but biologically implausible classifications ^48,49^. Our qPCR results also did not yield any positive Mpox results from environmental samples to confirm these sequencing results. However, continuing surveillance at these sites is still highly recommended. Taken together, these findings highlight the importance of confirming low-abundance metagenomic signals of high-consequence pathogens using independent analytical approaches before epidemiological interpretation, as reports of such pathogens may trigger extensive public health investigations, surveillance activities, and response measures. In parallel with the ML activities, the ODIN-Mpox consortium is conducting complementary analyses in conventional laboratory settings, and the results may provide additional insights into these findings.^50^.

Tiling amplicon sequencing approach was first developed for Zika virus, and later it was widely adopted for other viruses including SARS-CoV-2 during the COVID-19 pandemic ^51–54^. The protocol was mainly tailored to clinical samples where low viral reads posed challenges for virus characterization ^53^. This approach was also employed to detect genomic variant of SARS-CoV-2 in WW ^54^. Our previous study demonstrated the feasibility of this approach to characterize Mpox clade IIb in WW ^19^. This approach was subsequently validated with additional clades from WHO Biohub. Although this approach did not construct any phylogenetic trees from environmental samples, Mpox clade Ia was accurately identified in four out of five clinical samples. Viral DNA in WW have a greater risk of degrading due to exposure to environmental elements like heat, moisture and microbial activity. Consequently, whole genome sequencing using tiled amplicon sequencing approach for MPXV in WW samples will be much more challenging than clinical samples, especially in WW where MPXV particles are present at very low abundance. Even with clinical samples, this was quite challenging, demonstrated by the fact that two clinical samples required two rounds of PCR amplification, and one sample was negative for Mpox after two rounds. In addition, WW is a much more complex sample matrix than clinical samples, resulting in the possibility of off-target amplification which in turn reduces the chance to obtain a Mpox consensus genome ^54^.

The MLs offer a valuable platform for the rapid deployment of diagnostic resources during critical phases of infectious disease outbreaks. Most ML systems described in the literature have targeted individual patient diagnostics in outbreak response settings ^8,10^. Although ML concept is not new, several aspects in our approach set us apart from previous studies: the application of a ML for environmental pathogen surveillance; flexible and field-deployable modular lab workflows; the innovative integration of simplified bioinformatic pipelines and data visualization that can be performed on a single laptop. The capacity to facilitate on-site environmental surveillance could significantly enhance early pathogen detection and support swift local containment measures. The present deployment also prioritized population-level pathogen monitoring through environmental matrices, enabling detection of pathogens circulating below clinical detection thresholds and providing advance warning before outbreak case counts rise. This epidemiological distinction has practical implications for how ML results are interpreted and acted upon: environmental detections indicate exposure risk at the community level rather than confirmed infection in individuals.

The deployment was successfully completed without major issues; however, a few challenges were faced before and during the operation. The first challenge was reagent and equipment procurement. A great effort was put into this work so that reagents and consumables were sourced locally in Tanzania or through regional African suppliers. Minimizing reagent dependence on European sources ensures the practical feasibility of ML implementation in these countries. Although this deployment relied partly on European suppliers, comparable reagents are expected to be increasingly accessible through African distribution networks in future deployments. The second challenge was the hot weather. Several sequencing runs were abruptly terminated due to high temperatures that exceeded the operating limits of Nanopore sequencers. However, we discovered that the Mk1D device, the newest device from nanopore, performed much better under hot condition compared to the previous version Mk1B. Therefore, we recommend using Mk1D in such hot weather conditions. The third challenge we encountered was language barriers, as some researchers were from French-speaking countries (DRC and Burkina Faso). Although numerous AI-assisted tools were available for translation, having a native French-speaker in the team greatly improved communication and interaction with other researchers.

Environmental molecular signals from a single one-month deployment represent first-line screening rather than confirmatory diagnosis. Detection of pathogen-associated sequences in an environmental sample does not imply active human transmission; results require triangulation with clinical surveillance data, epidemiological context, and where relevant, targeted confirmatory assays (e.g. *ctxA* PCR for cholera, orthopoxvirus clade typing for Mpox). The design of combining metagenomic sequencing with qPCR assays allows more reliable results with high-confidence interpretation. Notably, for samples collected in Kinshasa, DRC, in the second half of 2025, environmental detection of *Vibrio cholerae* using qPCR and metagenomic methods during our ML deployment was observed during a period of active cholera transmission, supporting the potential value of environmental surveillance as a complementary tool for outbreak monitoring. Confirmation of toxigenic O1/O139 strains requires targeted PCR amplification of virulence genes such as *ctxA*. Therefore, Biomeme qPCR assays targeting a *V. cholerae* virulence gene (ctxA) were applied, providing an additional but distinct line of evidence to support metagenomic detections of *V. cholerae*. Nevertheless, the one-month deployment timeframe limits temporal conclusions, and the absence of replicate sampling at all sites reduces the ability to distinguish stable environmental signals from transient contamination events. Post-deployment activities for cross-method validation are still recommended in future operations, especially for high-consequence pathogen detections.

## 5 Conclusions

We demonstrated the successful deployment of a fully offline, modular environmental surveillance platform integrating mobile laboratory workflows, bioinformatic analysis, data visualization, and capacity building across multiple partner countries in sub-Saharan Africa. MLs and environmental monitoring have proven effective for rapid response to disease outbreaks, playing a crucial role in mitigating and controlling the spread of pathogenic threats. We demonstrated that a modular ML can deliver end-to-end molecular surveillance, from sampling to analysis, across diverse environmental matrices, with actionable turnaround times and embedded capacity transfer.

With respect to the three objectives guiding this deployment: 1. Autonomous operation: The mobile laboratory operated reliably for one month in an off-grid setting, powered by a gas generator and requiring no internet connectivity for any analytical workflow. All components fit within two vehicles and could be assembled or disassembled within approximately three hours. 2. Field data generation and visualization: Both targeted (qPCR) and untargeted (metagenomic) methods successfully generated environmental surveillance data on priority pathogens across three countries. The internet-independent Enlighten dashboard enabled on-site interpretation of results, with visualization templates updated remotely during the deployment. 3. Training and capacity transfer: Twenty-five researchers from four countries were trained across all laboratory modules, with several independently performing the full pipeline by the end of the deployment. Language barriers were overcome using AI-assisted protocol translation.

Future iterations of the mobile laboratory concept should aim for fully local reagent procurement through African distributors, permanent installation of field-ready systems at partner institutions, and expansion of the pathogen detection panel to include RNA viral targets, which were excluded from this deployment due to logistical constraints. The modular architecture demonstrated here provides a scalable foundation for building self-sustaining environmental surveillance capacity in resource-limited settings, with potential for rapid reconfiguration in response to emerging pathogen threats.

## Ethics approval

Ethical approvals were obtained from the relevant ethics review committees in participating countries (in the DRC: 585/CNES/BN/PMMF/2024 du 07/10/2024, in Tanzania: NIMR/HQ/R.8a/Vol.IX/4654; NIMR/HQ/R.8a/Vol.IX/5028, in Burkina Faso: Comité d’éthique pour la recherche en santé, N°2024-05-138). Clinical samples from patients in the DRC were anonymized prior to analysis, and no personal identifiers were accessible to researchers.

## Data Availability

All data produced in the present work are contained in the manuscript

https://www.ebi.ac.uk/ena/browser/view/PRJEB11861

## Acknowledgements

This study was supported by the European Commission H2020 and EDCTP3 through the ODIN project (grant no. 101103253, Strengthening Environmental Surveillance to Advance Public Health Action) and the ODIN-Mpox project (grant no. 101195186, Implementing wastewater and environmental surveillance for Mpox in sub-Saharan Africa). We would like to thank all participants from the partner organizations in Sub-Saharan Africa who took part in the training under both projects. Thank you for your active participation, fruitful collaboration, and valuable feedback on the ODIN Mobile Lab. We would like to thank the WHO BioHub System and the European Virus Archive Global (EVA GLOBAL) for sharing Mpox material including inactivated viruses and their DNA.

## Declaration on the use of AI

Artificial intelligence (AI) tools were used in the preparation of this manuscript to assist with text drafting, reference verification, and data summarisation. Specifically, GitHub Copilot (Microsoft/OpenAI) was used to generate and refine sections of the methods and results text, assisted with citation retrieval from DOI metadata, and contributed to the production of data analysis scripts used to generate the figures presented herein. All AI-generated content was reviewed, verified against primary data sources, and edited for accuracy and scientific appropriateness by the human authors. The authors take full responsibility for the integrity and accuracy of all content in this manuscript.

Large language models were not used to generate, interpret, or alter scientific data. AI tools did not participate in study design, sample collection, laboratory analyses, or any aspect of the fieldwork. The use of AI assistance in manuscript preparation is disclosed in accordance with the policies of the target journal.

## Author contributions

T.T. contributed to investigation, data curation, formal analysis, and writing of the original draft, as well as review and editing of the manuscript. J.C. contributed to investigation, data curation, formal analysis, visualization, writing of the original draft, and review and editing. D.S. contributed to investigation, data curation, formal analysis, visualization, writing of the original draft, and review and editing. A.B. contributed to data curation, formal analysis, and review and editing. G.F. contributed to data curation, formal analysis, visualization, and review and editing. A.L.T. contributed to environmental sampling, formal analysis, and review and editing. P.Mk., E.L., H.S., Ja.C., M.K., E.N., P.Mb., P.L., and B.K. contributed to environmental sampling, investigation, and review and editing. S.S. and E.S contributed to environmental sampling, investigation, and review and editing. V.B., M.C.T., and V.M.T. contributed to funding acquisition, local project activity coordination, sample provision, and review and editing. T.P. contributed to work package leadership, funding acquisition, and review and editing. R.L. contributed to overall project coordination, funding acquisition, and review and editing. A.K. contributed to conceptualization, funding acquisition, supervision, investigation, data curation, formal analysis, visualization and writing of the original draft, as well as review and editing of the manuscript.

