## Supplementary document 1 for "From sample to insight: Onsite training and implementing mobile environmental surveillance in sub-Saharan Africa"

### Supplementary document 1: **Testing tiling amplicon sequencing approach using WHO Mpox clades**

To facilitate the future processing and characterization of MPXV in the environmental samples, we conducted a series of experiments designed to demonstrate that our approach and protocol can be successfully applied to characterize MPXV strains belonging to different phylogenetic clusters, including clades Ia, Ib, and IIb.

Some aspects of these spiking experiments have been described in Bagi et al.,[1](2026). publication also provides detailed methodological information, including protocols for DNA extraction, library preparation, sequencing, and the bioinformatics pipeline used for data analysis.

The purpose of this add-on set-up was to test what the tool output would be if there were multiple Mpox strains present in the wastewater. Wastewater is a complex environmental sample – the presence of various strains in the sample is something should be expected.

The tool was mainly built to characterize Mpox strains from clinical samples; hence the input is normally only a single Mpox strain. The tool prepares a consensus sequence from MPXV genomes that have been DNA sequenced using a pooled tiling amplicon strategy. The inactivated virus strains were shipped to NORCE at Tromsø. Each strain was sent in two tubes.

| Reference number | Name | Variant | Volume per tube (Viral load PFU) |
|---|---|---|---|
| 2023-WHO-LS-008 | MPXV, strain hMpxV/DRCINRB/22MPX0422C/2023 | clade Ia | 0.9 ml (7.66E+04 PFU) |
| 2024-WHO-LS-002 | MPXV, strain hMpxV/DRCINRB/24MPX0205V/2024 | clade Ib | 0.9 ml (2.35E+05 PFU) |
| 2024-WHO-LS-011 | MPXV, strain hMpxV/CHHUG/38235549/2022 | clade IIb | 0.9ml (1.93E+05 PFU) |

Experiment set-up 1:

Sample 26: WW + 150 ul Mpox Ia

Sample 27: WW + 150 ul Mpox Ib

Sample 28: WW + 150 ul Mpox IIb

Sample 29: WW + 50 ul each Mpox Ia/Ib/IIb

Sample 30: WW only

Sample 31: WW + 100 ul Mpox IIb from Slovenia

[1] Bagi et al., 2026 - https://doi.org/10.64898/2026.04.01.26349919

1. Mix all Mpox clades and run with rapid sequencing kit (no barcodes) – mix 5ul each 26 to 29 -> total 20ul of the mix

In the first run, single strain-spiked and multiple strain-spiked DNA samples were added into the same tube and library-prepped before loading onto a Flongle flow cell.

The result shows that the consensus sequence was in Mpox clade IIb cluster in the phylogenetic tree but shifted a few nodes away from the location of used Mpox clade IIb strain if present alone.

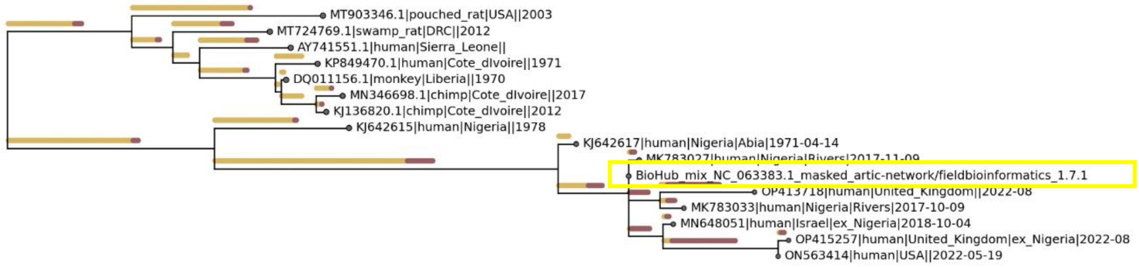


**Figure S1:** Mpox-spiked WW DNAs were mixed, followed by tiling amplicon sequencing protocol before being loaded on a flow cell without barcoding step

2. Barcode each clade and clade mix

Samples were prepared following the tiling amplicon sequencing and then barcoded separately before being sequenced on MinION platform.

The phylogenetic tree showed expected outcome for single Mpox strain samples: sample 27 (clade Ib) and sample 28&31 (cladeIIb) except for sample 26 (clade Ia)

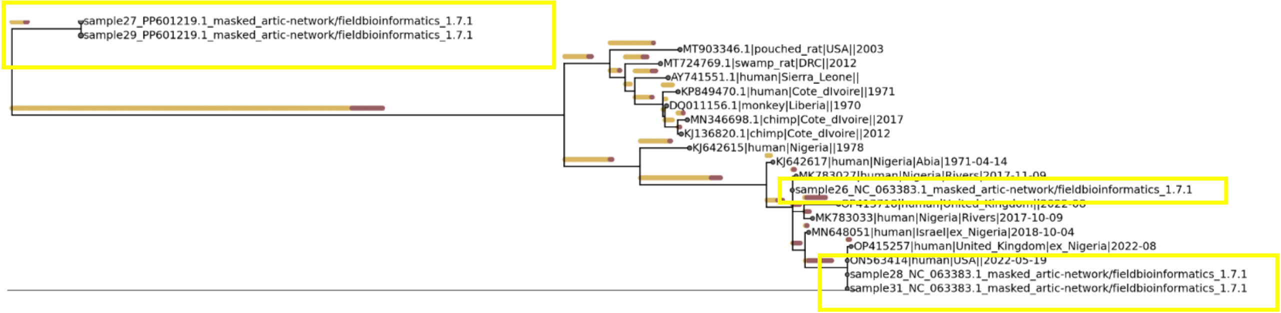


**Figure S2.** Mpox-spiked WW DNAs were amplified, pooled, and barcoded separately before being loaded on a flow cell. Clade Ib and clade IIb were accurately identified while clade Ia was mis-identified

Experiment set-up 2:

Sample 32: WW only

Sample 33: WW + 150 ul Mpox Ia (from tube 1)

Sample 34: WW + 150 ul Mpox Ia (from tube 2)

Sample 35: WW + 50 ul Mpox Ib + 100 ul Mpox IIb

Sample 36: WW + 100 ul Mpox Ib + 50 ul Mpox IIb

For this experiment, the second-round PCR was performed using first-round PCR product as a template. This is to amplify the number of amplicons. Mpox clade Ia was correctly identified in this set-up for both tube 1 and tube 2. Pre-extracted Dutch Mpox clade II after the second-round PCR was also mapped to the tree.

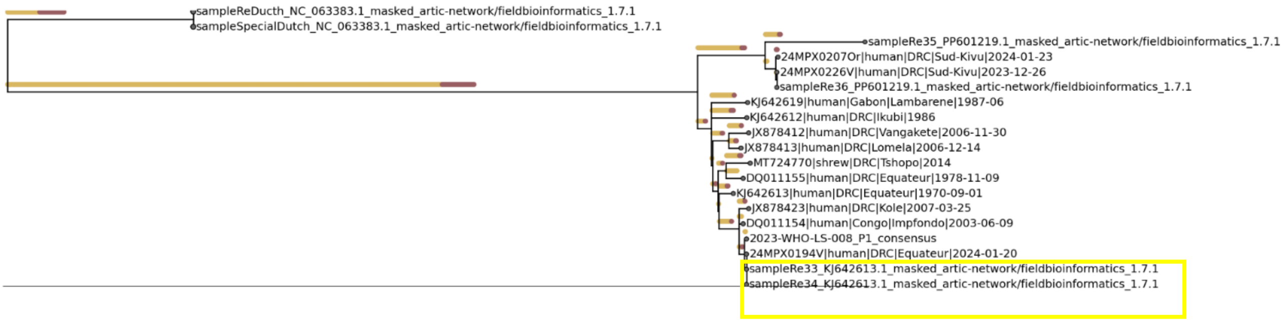


**Figure S3.** Clade Ia was accurately identified after two-round PCR amplification

#### Conclusion:

The tiling amplicon sequencing workflow and pipeline were able to identify all WHO Mpox clades (Ia, Ib, IIb). However, it may require extra PCR amplification step for an accurate identification if the starting material is insufficient for PCR in a single round.
